# Clinical Performance of SARS-CoV-2 Molecular Testing

**DOI:** 10.1101/2020.05.06.20093575

**Authors:** Daniel A. Green, Jason Zucker, Lars F. Westblade, Susan Whittier, Hanna Rennert, Priya Velu, Arryn Craney, Melissa Cushing, Dakai Liu, Magdalena Sobieszczyk, Amelia K. Boehme, Jorge L. Sepulveda

## Abstract

Molecular testing for severe acute respiratory syndrome coronavirus 2 (SARS-CoV-2) is the gold standard for diagnosis of coronavirus disease 2019 (COVID-19), but the test clinical performance is poorly understood. From 3/10/2020-5/1/2020 NewYork-Presbyterian laboratories performed 27,377 SARS-CoV-2 molecular assays from 22,338 patients. Repeat testing was performed in 3,432 patients, of which 2,413 had negative and 1,019 had positive first day results. Repeat-tested patients were more likely to be older, male, African-American or Hispanic, and to have severe disease. Among the patients with initially negative results, 18.6% became positive upon repeat-testing. Only 58.1% of any-time positive patients had a result of “detected” on the first test. The clinical sensitivity of COVID-19 molecular assays is estimated between 66.2 % and 95.6%, depending on the unknown number of false negative results in single-tested patients. Conversion to a negative result is unlikely to occur before 15 to 20 days after initial testing or 20-30 days after the onset of symptoms, with 50% conversion occurring at 28 days after initial testing. Forty-nine initially-positive patients converted to negative and then back to positive in subsequent days. Conversion from first day negative to positive results increased linearly with each day of testing, reaching 25% probability in 20 days. In summary, our study provides estimates of the clinical performance of SARS-CoV-2 molecular assays and suggests time frames for appropriate repeat testing, namely 15 to 20 days after a positive test and the same or next 2 days after a negative test in a patient with high suspicion for COVID-19.

## Introduction

Coronavirus disease 2019 (COVID-19) which is caused by severe acute respiratory syndrome coronavirus 2 (SARS-CoV-2) (1, 2) is a global pandemic with mortality significantly higher than seasonal influenza (3). The rapid sequencing of SARS-CoV-2 genomes (1, 2) has allowed the development of multiple real-time reverse transcription-polymerase chain reaction (RT-PCR) assays that have become the gold standard to detect viral RNA and identify patients with COVID-19 as well as asymptomatic carriers. However, some patients with positive chest radiologic findings and symptoms suspicious for COVID-19 have been reported to test negatively by SARS-CoV-2 RT-PCR and require multiple consecutive tests to convert to a positive result (4, 5). There is limited information on the clinical performance characteristics of the SARS-CoV-2 molecular tests in real world cases, namely what is the predictive value of a negative result in patients suspected of COVID-19 and what is the relationship between the course of the disease, viral shedding, and positivity of the various molecular assays. During the 2020 pandemic of COVID-19, we tested 27,377 samples from 22,338 patients with SARS-CoV-2 molecular assays performed at NewYork-Presbyterian (NYP) clinical laboratories. The assays used in this patient population included six platforms based on reverse-transcription and nucleic acid amplification to detect SARS-CoV-2 specific RNA sequences. Our goal is to identify the clinical performance characteristics of SARS-CoV-2 molecular assays for diagnosis and stratification of COVID-19 patients and determine the dynamics of SARS-CoV-2 molecular assay results over time in a large dataset from multiple hospital locations in the New York City area.

## Methods

### Laboratory Dataset

We extracted records of all the SARS-CoV-2 tests performed at NYP laboratories from our laboratory information system (Cerner Millennium, Cerner Corporation, North Kansas City, MO) using a custom Cerner Command Language query. We tested a total number of 22,338 patients between March 10, 2020 and May 1, 2020 for SARS-CoV-2 by molecular assays at NewYork-Presbyterian affiliated hospitals and performed a total of 27,377 assays on nasopharyngeal (initially also on oropharyngeal) swab samples. The vast majority of the SARS-CoV-2 tests were reverse-transcription real time PCR assays performed with the high-throughput automated cobas 6800 (Roche Molecular Systems, Branchburg, NJ) platform (N=19,195), which went live at Columbia-NYP laboratory on 3/15/2020 and at Cornell-NYP laboratory on 3/30/2020. A smaller number of assays were performed since 3/10/2020 with in-house developed tests under FDA Emergency Use Authorization using the Rotor-Gene Q (N=1795, Qiagen, Valencia, CA) and 7500 Fast (N=89, Thermo-Fisher Scientific, Waltham, MA) instruments. From 4/3/2020 thereafter other platforms were introduced in the NYP laboratories, including the Xpert Xpress SARS-CoV-2 test analyzed on the GeneXpert (N=265) and Infinity (N=5954) platforms (Cepheid, Sunnyvale, CA), the ID Now (N=53), (Abbott, Chicago, IL) and the Panther Fusion (N=26) (Hologic Inc., San Diego, CA) instruments. Commercial molecular viral assays were validated and performed at NYP Hospital Laboratories following manufacturers’ recommendations.

### Patients

The NYP laboratory SARS-CoV-2 testing dataset contains basic demographic information on each patient, including age, gender, race, and health care encounter type and location at the time of order. To better understand SARS-CoV2-2 test utilization and investigate the association with clinical information to the SARS-CoV-2 test results on a subset of patients, we merged our laboratory dataset with the Columbia COVID-19 CARE clinical database.

The COVID-CARE clinical database is an interdisciplinary database, managed by the Columbia Division of Infectious Diseases and contains data on all patients tested for SARS-CoV-2 at the New York Presbyterian West Campus (Milstein Hospital, Allen Hospital, Morgan Stanley Children’s Hospital). This interdisciplinary database contains the work of numerous divisions within the Departments of Medicine, Pediatrics, Neurology, and Obstetrics and Gynecology. The database is comprised of a general primary instrument and multiple sub-specialty instruments resulting in over 720 curated fields per patient. Data was extracted from the medical record through manual review by a team of medical students and sub-specialty fellows and attendings. Manually curated data is combined with structured data from the electronic medical records, pharmacy, and laboratory systems to complete the dataset. As of May 1^st^ 2020, 1,624 patients had undergone manual review (Supplementary Table 1).

Clinical factors and other patient characteristic are compared between SARS-CoV-2 positive and SARS-CoV-2 negative patients in Supplementary Table 2, and between repeat-tested and single-tested patients in Supplementary Table 3. This study was approved by the Institutional Review Boards of Columbia and Cornell Universities.

### Data Analysis

Categorical data was presented as frequency and percent. Continuous data was presented as mean (standard deviation) or median and range. Differences between groups were compared using Chi-square tests for categorical variables, and linear ANOVA for continuous variables. Correction for multiple testing was performed using the false discovery rate method of Benjamini-Hochberg (6). Confidence intervals for sensitivity and negative predictive values were calculated using the *epiR* R package. Time to event using Kaplan Meier curves were used to investigate time to conversion from initially negative to positive and from initially positive to negative. All statistical analyses were performed using the R statistical language (7).

## Results

A total of 18,906 assays were performed once per patient and 8,471 assays were performed in 3,432 (15.4%) patients with more than one test completed in the span of 1 to 49 (median = 8) days between assays. There were 2,280 patients with multiple SARS-CoV-2 molecular assays performed who had a first negative, invalid, or indeterminate result and 797 patients who had an initial positive result.

### Demographic and clinical characteristics

Patients with any positive SARS-CoV-2 result differed from those that were never positive for SARS-CoV-2 even with repeat testing (Supplementary Table 2). Notably, SARS-CoV-2 positive patients were more likely to be male, older than 44 years, and of self-reported African-American or Hispanic / Latino ethnicity and less likely to be Asian or Caucasian (Supplementary Table 2, all p<0.001). Not surprisingly, severe disease was more likely in SARS-CoV-2 positive patients, as indicated by a higher fatality rate (6.2% vs. 3.8%, p=0.048), presence of symptoms (91.8% vs. 72.4%, p=0.004), need for intubation (10.7% vs. 7%, p=0.026) and frequency of decompensation, as defined by an outcome of intubation, death, or discharge to hospice (13.4% vs. 8.4%, p=0.005).

To determine which factors were associated with repeat testing for SARS-CoV-2 we analyzed demographic and clinical features of repeat-tested as compared to single-tested patients (Supplementary Table 3). Our data show higher age (median = 59.9 vs. 53.4, p<0.001), higher frequency of male gender (52.2% vs. 44.3%, p<0.001), and different distribution of self-reported race and ethnicity in repeat-tested as compared to single-tested patients, with African-Americans and Hispanics / Latinos being more likely to be repeat-tested (p<0.001). At the time of the first test order, admitted patients were significantly more represented in the repeat-tested population in contrast to patients visiting the emergency department and outpatients (p<0.001). Compared with single-tested patients, repeat-tested patients were 3.2 times more likely to be admitted to the ICU (29.7% vs. 9.4%, p<0.001), 4 times more likely to be intubated (24.4% vs. 6.1%, p<0.001), 3.4 times more likely to decompensate (27.6% vs. 8.1%, p<0.001) and 1.7 times more likely to die during the observation period (8% vs. 4.7%, p=0.038).

### SARS-CoV-2 Test Performance

The characteristics of the SARS-CoV-2 tests performed in repeat-tested compared to single-tested patients are described in Supplementary Table 4. The vast majority of tests were performed with the cobas 6800 and the use of the various assays was not significantly different between repeat and single-tested patients, except for a small number of patients initially tested with the Thermo-Fisher 7500 assay, which was more likely in repeat-tested patients (p<0.001, standardized Pearson residuals for repeat-tested patients = 3.9).

Among all the repeat-tested patients, 23.2% were positive on the first test (26.5% when indeterminate results were included). If a negative test was repeated on the first day, the positivity rate increased to 26.4% (29.7% with indeterminate results). Overall positivity among repeat-tested patients over the course of the study period was 39.9% in contrast to 49% for single-tested patients (p<0.001). When indeterminate results were counted as positive, 42.9% of repeat-tested patients were positive over the course of the study period in contrast to 50% of single-tested patients (p<0.001).

Indeterminate results are generally considered presumptive positive and occur when only one of two molecular targets is detected. In our repeat-tested patients with an initial result of “Indeterminate”, 54.4% ultimately had a result of “Detected”, as compared to 7% that remained indeterminate upon repeat testing and 38.6% that converted to a negative status during our study period, thus suggesting that it is acceptable to consider these patients positive. However, it is unclear whether the initially indeterminate patients that converted to negative were false positives or presented with low viral loads.

In contrast, a result of “Invalid” reflects the failure to amplify the built-in control and is likely related to poor sampling or inadequate RNA extraction usually due to high viscosity of the samples. In our study of repeat test patients, 52.7% ultimately became positive, 1.2% repeated as “Indeterminate” and 45.6% repeated as ‘Not Detected’. For the analysis of clinical sensitivity of SARS-CoV-2 molecular tests, we considered patients with invalid results as “negative”, as these results can be considered clinically false negatives in the sense that the patient may be infected and the test failed to yield a positive result. In practice, invalid results should always be repeated, preferably with a new sample, as the results are unpredictable.

Initial negative, invalid, or indeterminate SARS-CoV-2 test results were much more frequent among repeat-tested patients (Supplementary Table 4, p<0.001) and conversely, patients without a positive initial result were more likely to be repeated (21%) than initially positive patients (8%, p<0.001).

A subset of the cobas 6800 tests (N=5,343) had cycle threshold (Ct) values available for analysis. The Ct represents the PCR cycle, interpolated to two decimal digits, at which the real-time fluorescent signal crosses a pre-defined threshold for positivity. The Ct is inversely proportional to the concentration of viral RNA. The cobas SARS-CoV-2 RT-PCR assay amplifies two specific targets: Target 1 is located in the ORF1ab non-structural region that is unique to SARS-CoV-2 and Target 2 is a conserved region of the structural protein envelope E-gene common to all members of the Sarbecovirus sub-genus of coronavirus, which include SARS-CoV-2 and SARS-CoV (8, 9). The cobas assay also includes an internal control for assay performance that has no homology to the coronaviruses.

When comparing positive and indeterminate results from repeat-tested with single tested patients, there were no differences in indeterminate Ct values, which is expected as by definition the indeterminate results represent high Ct values. In contrast, the repeat-tested group (N=795) had significantly higher target 2 Ct values (median = 29.1 vs. 27.3, p<0.001 and frequency of target 2 Ct’s above 30 (45.8% vs. 36.9%, p<0.001) compared to single-tested patients (N=4,548), indicating lower viral load in the repeat-tested samples (Figure 1 and Supplementary Table 4).

**Figure 1:**
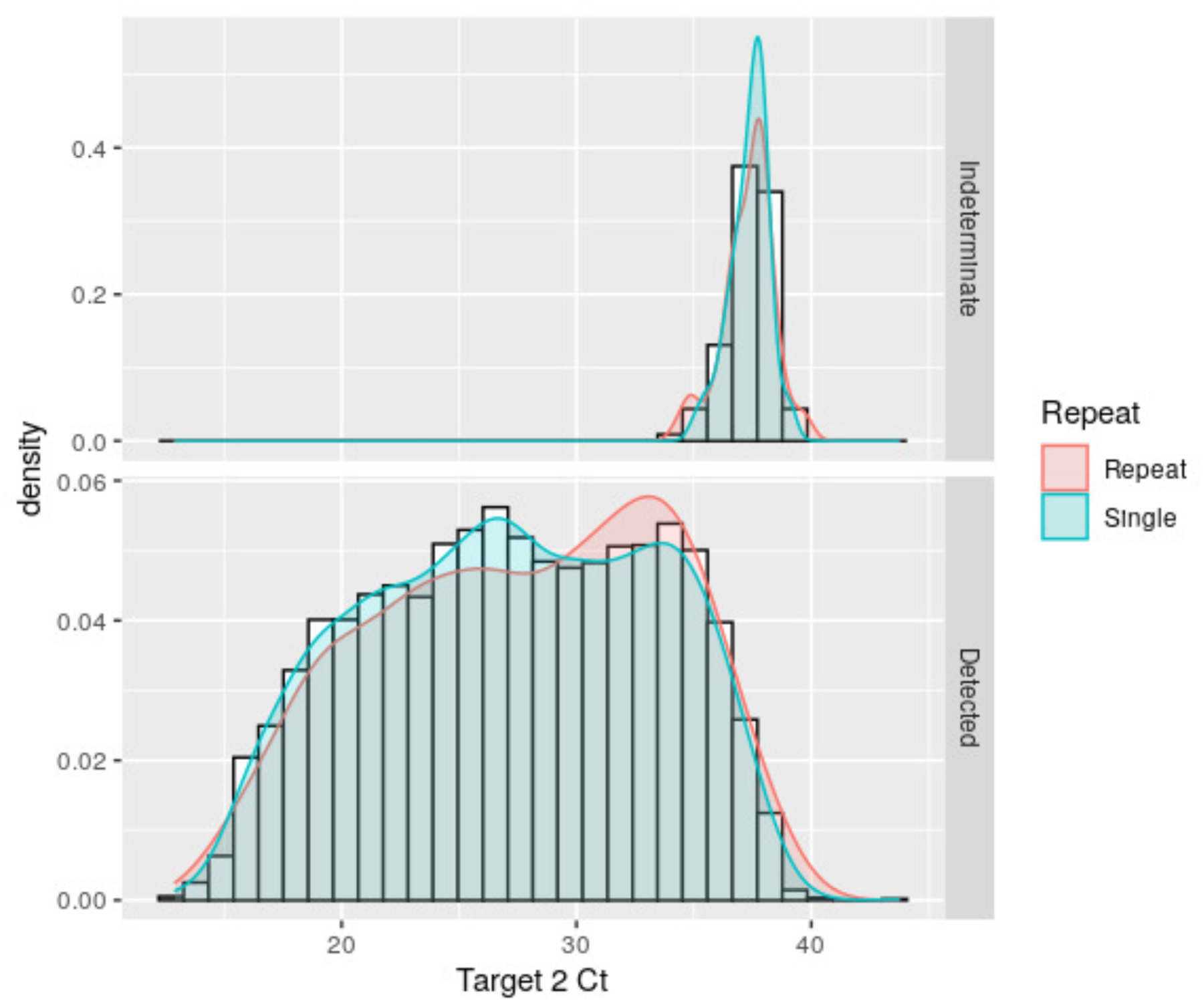
Density distribution of cobas SARS-2-CoV2 Target 2 Ct in repeat-tested vs. single-tested patients. Top panel: Ct values from results reported as “intermediate”; bottom panel: Ct values from results reported as “detected”.

### Analysis of conversion rates of repeat-tested patients

In this study, we classified repeat-tested patients in two groups according to their initial SARS-CoV-2 results: those that were initially positive, for whom repeat testing was most likely intended to ascertain recovery and non-infectiousness, and those that had an initial result of negative, indeterminate, or invalid, in whom persistent clinical suspicion for COVID-19 likely motivated repeat ordering of the test. Supplementary Table 4 shows the different clinical characteristics between repeat-tested and single-tested patients.

For the time-dependent analysis of conversion rates, we considered “initially positive” patients with any “Detected” or “Indeterminate” SARS-CoV-2 result obtained during the first calendar day of testing rather than the first positive test, to reduce bias due to nasopharyngeal sampling inadequacy (Table 1). Conversely, patients without a result of “Detected” or ‘Indeterminate” in the first day were labeled as “initially negative”. Among the 2,413 initially negative repeat-tested patients, 18.6% became positive upon repeat testing on subsequent days (Supplementary Table 5), indicating a negative predictive value of 81.3% (95% CI = 79.7 to 82.8) in this repeat-tested population with the prevalence present at the time of the study.

**Table 1:** Number of SARS-CoV-2 molecular test results over the course of repeat testing, grouped by the highest test result on day 1.

| **Highest Result** | Invalid | Not Detected | Indeterminate | Detected | Total |
| --- | --- | --- | --- | --- | --- |
| **Any Day —>** | (N=1) | (N=1960) | (N=100) | (N=1371) | (N=3432) |
| **Highest Day 1** | p value <0.001<sup>1</sup> |  |  |  |  |
| **Result** |  |  |  |  |  |
| Invalid | 1 (100%) | 80 (4.1%) | 3 (3.0%) | 79 (5.8%) | 163 (4.7%) |
| Not Detected | 0 (0.0%) | 1880 (95.9%) | 35 (35.0%) | 335 (24.4%) | 2250 (65.6%) |
| Indeterminate | 0 (0.0%) | 0 (0.0%) | 62 (62.0%) | 50 (3.6%) | 112 (3.3%) |
| Detected | 0 (0.0%) | 0 (0.0%) | 0 (0.0%) | 907 (66.2%) | 907 (26.4%) |

1. Pearson’s Chi-squared test (adjusted for multiple comparisons)

In a separate analysis, we compared the results of the first test with the results from tests repeated the same day (Supplementary Table 5) or repeated any time after the first result (Supplementary Table 6). Among the patients with repeat testing who had initial results of “Invalid” (241), “Not Detected” (2280), or “Indeterminate” (114), 4.2% had ‘Detected’ results upon repeat testing in the first day of testing (Supplementary Table 5). This increase in positivity rate most likely results from a false-negative initial test due to pre-analytic factors such as sample inadequacy, incorrect swabbing technique, or stochastic sampling bias from low viral loads in the patient nasopharynx. After the first day of testing, repeating invalid, negative, or indeterminate results on the same day resulted in about 0.8% of additional positive results per day, for a total of 18.4% positives that were missed by the first test.

Among the 1,371 repeat-tested patients with one or more SARS-CoV-2 results of “Detected”, which can be assumed to be truly infected, only 58.1% were resulted as “Detected” on the initial test (Supplementary Table 5), and only 66.2% were reported as “Detected” on the first day (Table 1). Considering ‘Detected’ and “Indeterminate” as positive, 1,471 repeat-tested patients had one or more SARS-CoV-2 positive results over time; only 61.9% were positive on the initial test and only 69.3% had a positive result on the first day (Table 1). These data provide an estimate of the clinical sensitivity of the assay in the repeat-tested population, and establish a baseline to look at conversion rates from negative to positive.

Table 2A shows the number of patients who had an initial result of “Not Detected” on day 1 who converted to a SARS-CoV-2 positive status grouped per time after the initial test. Conversely, Table 2B shows rates of conversion to negative for patients with a status of “Detected” on day 1. Supplementary Figure 1 shows the distribution per day after onset of symptoms (**a** and **b**) or after initial testing (**c** to **f**) of conversions from positive to negative (left-side) and from negative to positive (right-side) in the two groups of repeat-tested patients. For this analysis, positive status included “Detected” and “Indeterminate” and negative status included results of “Not Detected” with “Invalid” results excluded. Among the initially positive patients, the unadjusted distributions show a peak of conversion to negative between 30 and 40 days after symptoms or around 20 days after initial testing. Less than 10% of the patients who converted to negative converted before 15 days after onset of symptoms or 10 days after initial testing. In contrast, among the patients with initially negative results who converted to positive, most conversions occurred 10 days or less after onset of symptoms, and in the first 1-3 days after initial testing.

**Table 2:**
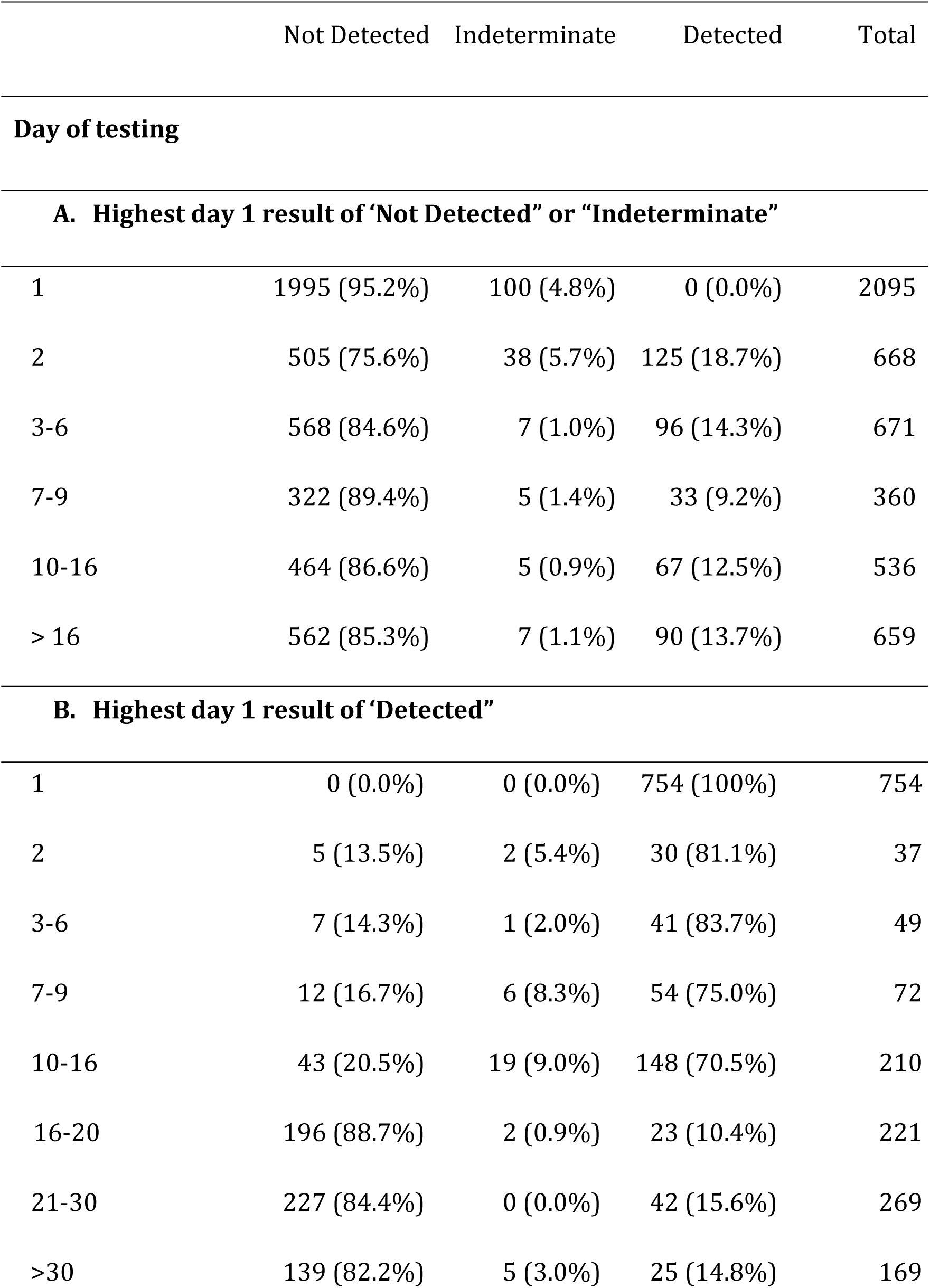
Distribution of repeat tests per day after a first day result of “Not Detected” or “Indeterminate” (A) or after a first day result of “Detected” (B).

|  | Not Detected | Indeterminate | Detected | Total |
| --- | --- | --- | --- | --- |
| **Day of testing** |  |  |  |  |
| **A. Highest day 1 result of ‘Not Detected’ or ‘Indeterminate’** |  |  |  |  |
| 1 | 1995 (95.2%) | 100 (4.8%) | 0 (0.0%) | 2095 |
| 2 | 505 (75.6%) | 38 (5.7%) | 125 (18.7%) | 668 |
| 3-6 | 568 (84.6%) | 7 (1.0%) | 96 (14.3%) | 671 |
| 7-9 | 322 (89.4%) | 5 (1.4%) | 33 (9.2%) | 360 |
| 10-16 | 464 (86.6%) | 5 (0.9%) | 67 (12.5%) | 536 |
| > 16 | 562 (85.3%) | 7 (1.1%) | 90 (13.7%) | 659 |
| **B. Highest day 1 result of ‘Detected’** |  |  |  |  |
| 1 | 0 (0.0%) | 0 (0.0%) | 754 (100%) | 754 |
| 2 | 5 (13.5%) | 2 (5.4%) | 30 (81.1%) | 37 |
| 3-6 | 7 (14.3%) | 1 (2.0%) | 41 (83.7%) | 49 |
| 7-9 | 12 (16.7%) | 6 (8.3%) | 54 (75.0%) | 72 |
| 10-16 | 43 (20.5%) | 19 (9.0%) | 148 (70.5%) | 210 |
| 16-20 | 196 (88.7%) | 2 (0.9%) | 23 (10.4%) | 221 |
| 21-30 | 227 (84.4%) | 0 (0.0%) | 42 (15.6%) | 269 |
| >30 | 139 (82.2%) | 5 (3.0%) | 25 (14.8%) | 169 |

Since we cannot be certain about the conversion rates due to a significant proportion of repeat-test patients having insufficient testing performed to detect conversion (right-censoring) we used a Kaplan-Meier approach to estimate the conversion rate by day of testing (Figures 2, 3 and Supplementary Figure 1 **e** to **h**) with the following assumptions:

**Figure 2:**
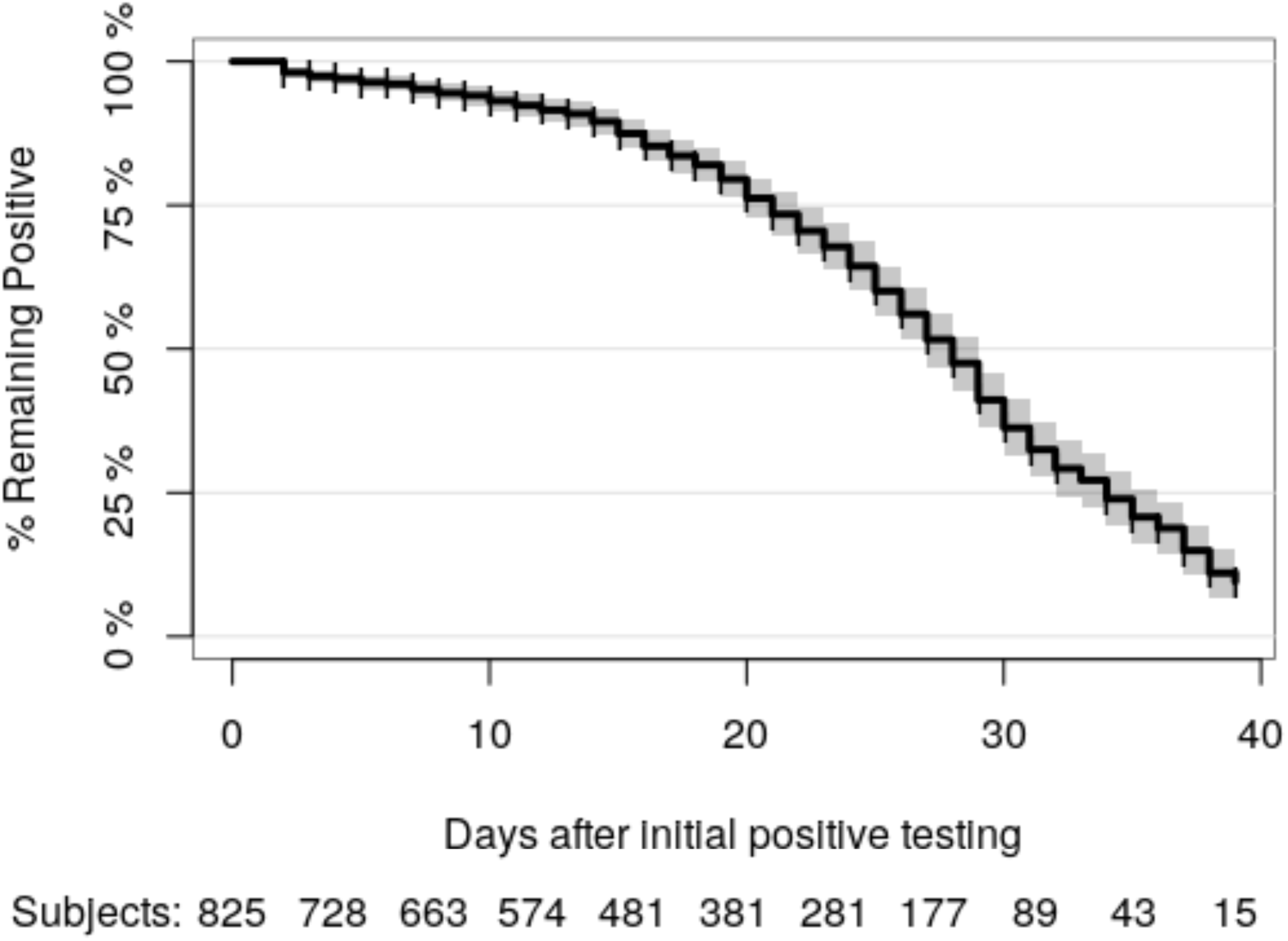
Kaplan-Meier estimate of conversion from an initially positive SARS-CoV-2 status on day 1 to a subsequent negative result.

**Figure 3:**
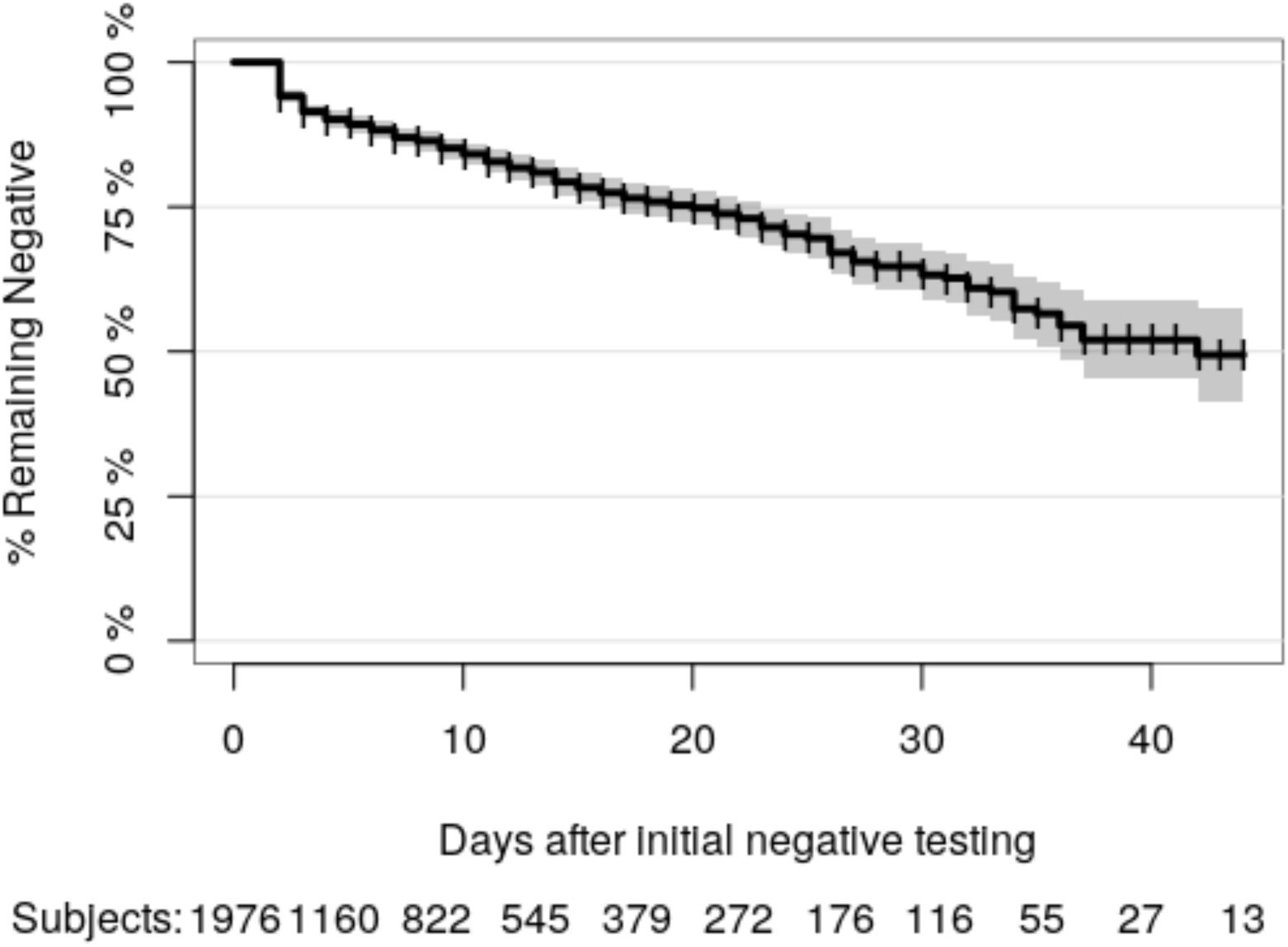
Kaplan-Meier estimate of conversion rate from initially negative SARS-CoV-2 status on day 1 to a subsequent positive result.

1. When there were multiple tests performed per patient in one day, the results were aggregated to the highest result, i.e. Detected > Indeterminate > Not Detected.
2. For initially positive patients, an event is defined as highest result in a day of “Not Detected”.
3. Conversely, an event is defined as a conversion from a “Not Detected”, or “Invalid” result at day 1 to “Detected” or “Indeterminate” on subsequent days.
4. If the last test result was unchanged relative to the first day result, the patient was considered right-censored at that time.
5. Only the first day and either the censoring day or the event day were used for each patient and indeterminate results were ignored.

The results show that the probability of converting from positive to negative in the initially positive, repeat-tested population is minimal until about 15 to 20 days after initial testing and reaches 50% at 28 days (95% CI = 27 to 29 days, Figure 2). In the initially negative repeat-tested population, the risk linearly increases every day with a 25% probability of conversion to positive of 20 days (95% CI = 16 to 23 days, Figure 3).

Since lack of conversion could be due to death of the patient, which occurred in about 8% of the repeat-tested population, we performed a competing risk analysis with death as the alternative event, using the *timereg* R package (10), and did not find significant differences in either the positive to negative or the negative to positive Kaplan-Meier cumulative probability of conversion (results not shown).

Interestingly, among the repeat-tested patients we have identified 11 patients that converted from an initial SARS-CoV-2 negative result to positive and back to negative over several days. We also identified 49 initially positive patients that became negative and later turned positive again. The time course of SARS-CoV-2 test results for these patients are shown on Supplementary Figure 2.

## Discussion

Our results from repeat-tested patients can be used to estimate the clinical sensitivity of the SARS-CoV-2 molecular testing in the population of patients that were selected for repeat testing, in contrast to the general population of tested patients. With all likelihood, most repeat testing on initial negative patients was performed either to follow a history of exposure, when the clinical profile did not fit the initial results, or when clinical presentation deteriorated after the initial result. Consistent with this hypothesis, repeat-tested patients were more likely to be older, male and of non-Caucasian race than single tested patients (Supplementary Table 3), consistent with the demographics of COVID-19 positive patients (Supplementary Table 2). Importantly, repeat-tested patients had worse outcomes as demonstrated by higher rates of decompensation, intubation, and mortality. Interestingly, repeat-tested patient who converted from positive to negative tended to be younger and present as outpatients, as compared to patients that remained positive, but there were no significant differences in clinical outcomes (Supplementary Table 7). In contrast, repeat-tested patients that remained negative tended to be inpatients, have longer admission duration, and were more likely to be extubated than those who converted to positive (Supplementary Table 8), suggesting that a significant number of repeat-testing in SARS-CoV-2 negative patients was performed in inpatients admitted before the pandemic or for non-COVID-19 reasons. Indeed, repeat-tested patients admitted before March 1st, 2020 were much more likely to have a persistent negative result (71.4%) than those admitted after March 1st, 2020 (39.7%, p<0.001).

In the absence of a more sensitive gold-standard, the repeat-tested patients with an eventual positive result can be considered true positives, as the analytical specificity of molecular testing is very high (3, 8, 9, 11, 12). It may be tempting to add all the 9,272 single-tested positive patients to the 1,371 repeat-tested positive patients to determine clinical sensitivity. However, we don’t know how many of the “Not detected” single-tested patients are false negatives, especially given the high frequency of asymptomatic or mildly symptomatic COVID-19 patients (13-15). Therefore, considering only the positive patients will inflate the estimated clinical sensitivity. Nevertheless, if we consider all negative results (repeated or not) to be true negative (i.e. a specificity of 100%) we can estimate the upper bound of the clinical sensitivity of a first initial result to be 94.6% (95% CI = 94.2-95%). If the test is repeated on the first day to account for nasopharyngeal sampling inadequacy, the upper boundary of the estimated sensitivity with these assumptions would be 95.6% (95% CI=95.2-96%). A lower bound of clinical sensitivity can be estimated by considering the worst case scenario that the same percentage of false negatives identified in repeat-tested patients would apply to the general population. With these assumptions, the clinical sensitivity of the SARS-CoV-2 assay can be estimated to be between 66.2% (95% CI = 63.6 to 68.7) and the above best case estimate of 95.6%.

The lower clinical sensitivity of the first day results in the ultimately positive repeat-tested patients suggests several possibilities:

1. Viral shedding increases over time in a recently infected patient and will eventually cross the detection threshold in subsequent samples. This possibility is suggested by the lower Ct values for target 1 and particularly target 2 on the repeat-tested patients (Supplementary Table 4 and Figure 1).
2. A significant number of samples are improperly collected, and repeat testing increases the probability of detection; this is particularly likely in the first day of testing, when suspicion may be high but the initial test results were negative or inconclusive, as shown in Supplementary Table 5.
3. Initially tested patients were truly negative and acquired the infection nosocomially after admission.

The mean interval between an initial negative test result and the first positive in patients who converted in our study was 9.4 days (95% CI = 8.4 to 10.5 days, N = 335), which is longer than reported by Ai et al, who showed a mean interval time between initial negative to positive RT-PCR results of 5.1 ± 1.5 days with a median of 4 days (N = 15) (4). We have insufficient data to calculate incubation time as only 1.3% of the patients had symptoms after the first test, with a median time between symptoms and testing of 1.3 days. The median interval between start of symptoms and the first test in our study of 4.8 days (95% CI = 4.5 to 5.4 days) is in line with the data from Lauer et al., who calculated a median incubation time of 5.1 days (95% CI = 4.5 to 5.8 days) in patients from China (16).

The third possibility of hospital-acquired infection explaining a low rate of initial true negative result is less likely because a) patients who converted from negative to positive had shorter intervals between admission and the first test and shorter duration of admission than those who remained negative (Supplementary Table 9 and b) there were no significant differences between repeat-tested inpatients and outpatients in the average time interval between the initial negative test and the first positive test (results not shown). If there were a large number of inpatients acquiring the infection in the hospital one would expect longer intervals between admission and positivity due to the cohort of patients already admitted before the pandemic, as compared to patients recently admitted with COVID-19. Rather, these data suggest a pattern of repeated ordering in uninfected inpatients with a lower likelihood of conversion.

Our analysis of repeat-tested patients with an initial positive result, using the Kaplan-Meier estimator, indicates that conversion to a negative result is unlikely to occur until about 15 to 20 days after initial testing or 20 to 30 days after start of symptoms, when the odds ratio significantly increase (Figure 2 and Supplementary Figure 1). Conversely, when a patient is suspected of COVID-19 but the initial test is negative, repeating the test steadily increases the probably of conversion to ‘Detected’ every day (Figure 3). Repeat testing is especially likely to yield a positive result if the initial test is indeterminate or invalid. For indeterminate results, this likely reflects low levels of the virus in the sample or low viral shedding in the nasopharynx early in the course of the infection. For invalid results, this likely reflects sampling inadequacy, often due to excess mucous in the sample. Our finding of significantly higher Ct in repeat-tested patients as compared to single-tested patients supports this hypothesis.

### Limitations of the study

This is an observational study without selection bias for the laboratory data, as all results were included in the analysis. However, the clinical data was restricted to a subset of patients seen at Columbia University Irving Medical Center campuses. Nevertheless, rates of positivity and demographic variables captured in the laboratory dataset were not significantly different between the other campuses, suggesting that the population in the clinical dataset is generally representative of the New York City patients tested for COVID-19. Other limitations of the study of repeat-tested patients include: 1) only 15% of the total patient’s tested had repeat testing done and 2) ordering of repeat testing was at the discretion of the health care providers and not performed according to a standard protocol, although test ordering was mostly accomplished through standardized electronic medical record order sets.

## Conclusions

Our data suggest that patients with a high clinical suspicion or exposure setting suggestive of COVID-19 should be tested by a molecular SARS-CoV-2 assay and it is appropriate to repeat the test the same day or in subsequent days if the results are initially negative. Conversely, for patients with a positive SARS-CoV-2 molecular assay result, repeating the test before at least 15 days after the first test is unlikely to yield a negative result. Whether repeat positivity represents active infection or detection of nonviable viral RNA is unknown. Further studies are needed to develop predictive models of the course and outcomes of COVID-19 using well-curated demographic, clinical, and laboratory datasets.

## Data Availability

The anonymized data stripped of personal health identifiers sufficient for reproducing the results in this manuscript will be shared upon request and review by NYP, Cornell and Columbia Universities.

## Acknowledgments

We would like to acknowledge the contributions of all the dedicated medical technologists and laboratory technicians who performed testing at NYP laboratories, the efforts of the NYP Laboratory Information Team, in particular Kelvin Espinal, Sarah Russell, Dennis Camp, Yingzhe Kuang, and Bulent Oral for designing queries and providing data extracts, and the volunteers who helped gather data from electronic medical records for the COVID-19 CARE database.

---

**Supplementary Figure 1:**
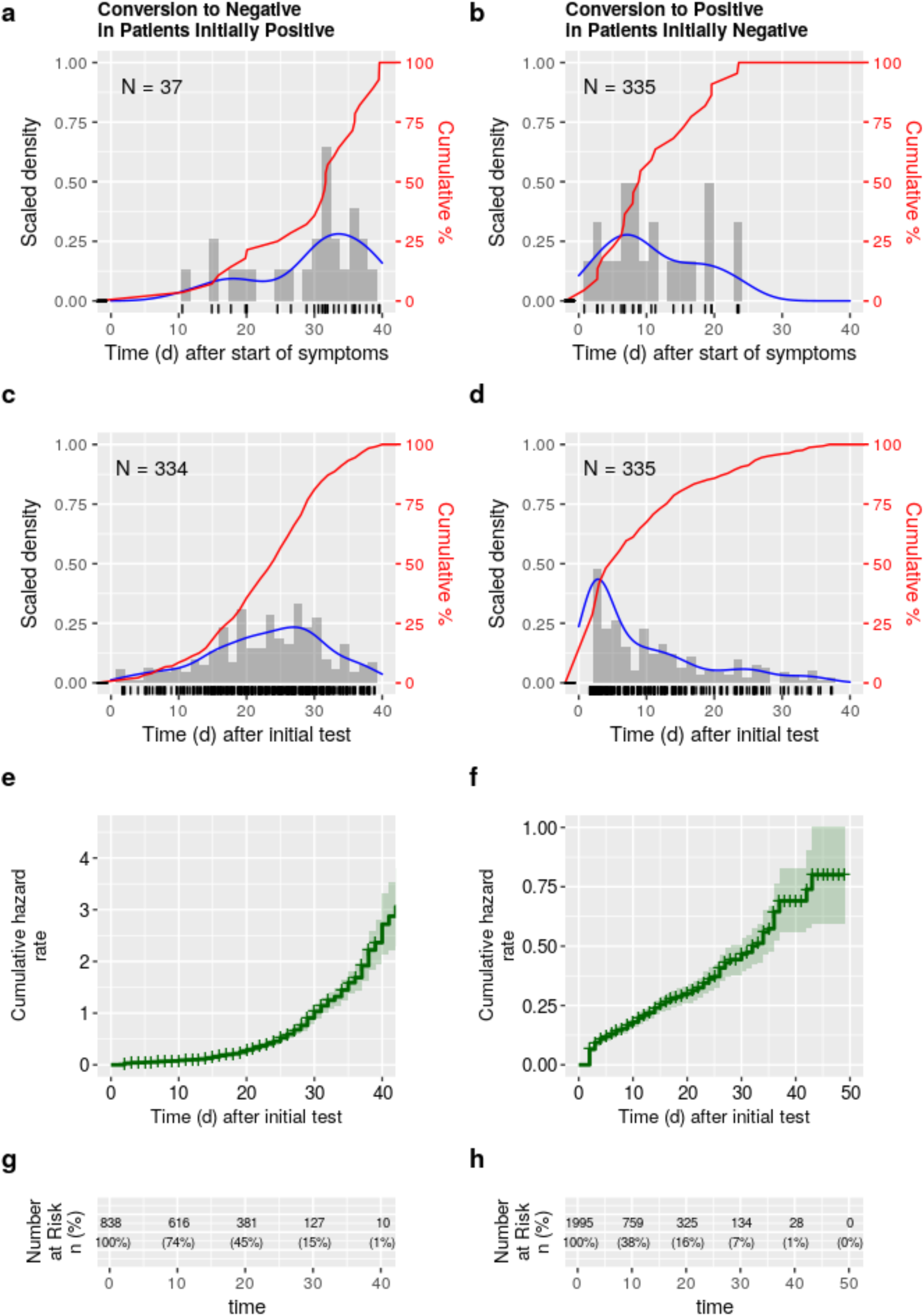
Conversion distributions for repeat-tested patients with initial SARS-CoV-2 negative (**a**, **c**, **e**, and **g**) and positive (**b**, **d**, **f**, and **h**) results. For the analyses in **a** to **d** only the first conversion events (i.e. negative to positive - **a** and **c** - or positive to negative - **b** and **d**) were considered. **N** represents the total numbers of patients analyzed. The distributions in **a** to **d** are represented by density-adjusted histograms (in grey bars), kernel probability density lines (in blue), unadjusted cumulative distributions (red line) and Kaplan-Meier hazard rates (green); the timing of individual results are represented by the black marks along the x-axis. For the Kaplan Meier estimated hazard rates (**e** to **h**), in addition to the conversion events, patients were considered censored at the time of the last unchanged result were included and the number of patients at risk for conversion at each time point are shown in **g** and **h**.

**Supplementary Figure 2:**
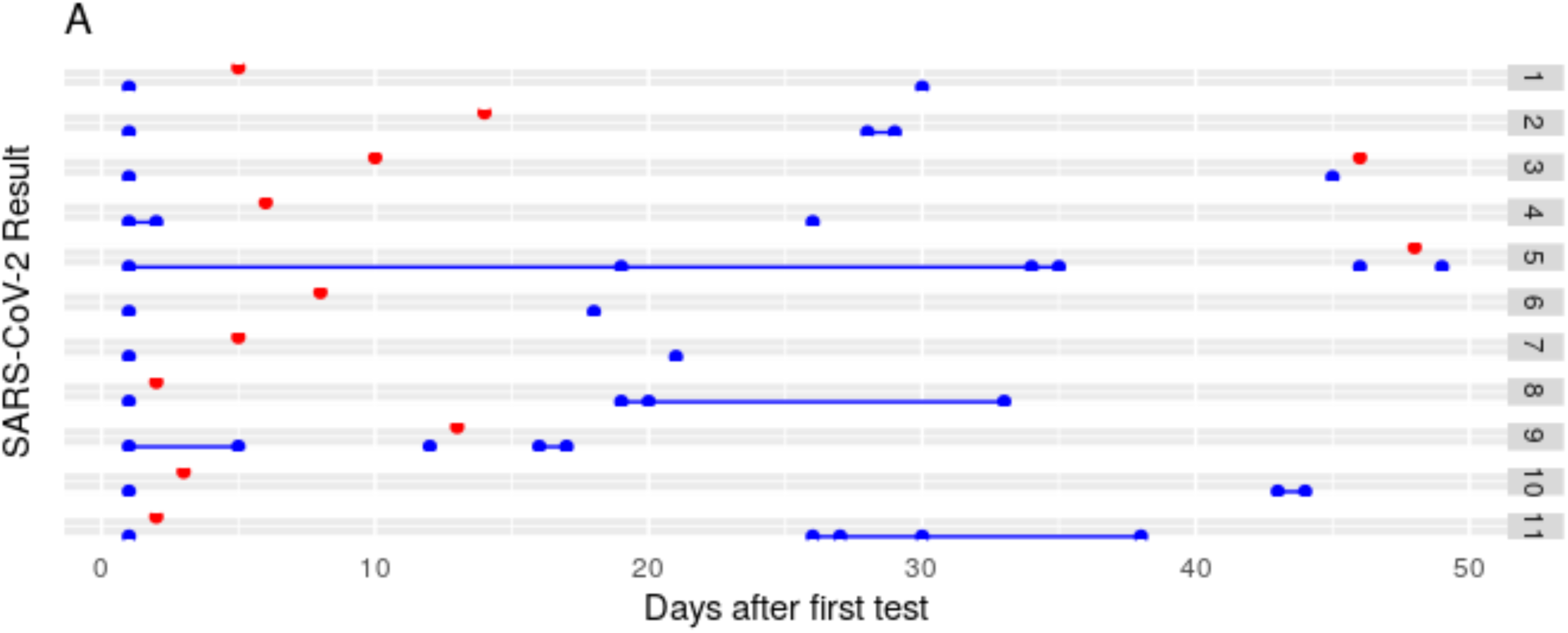
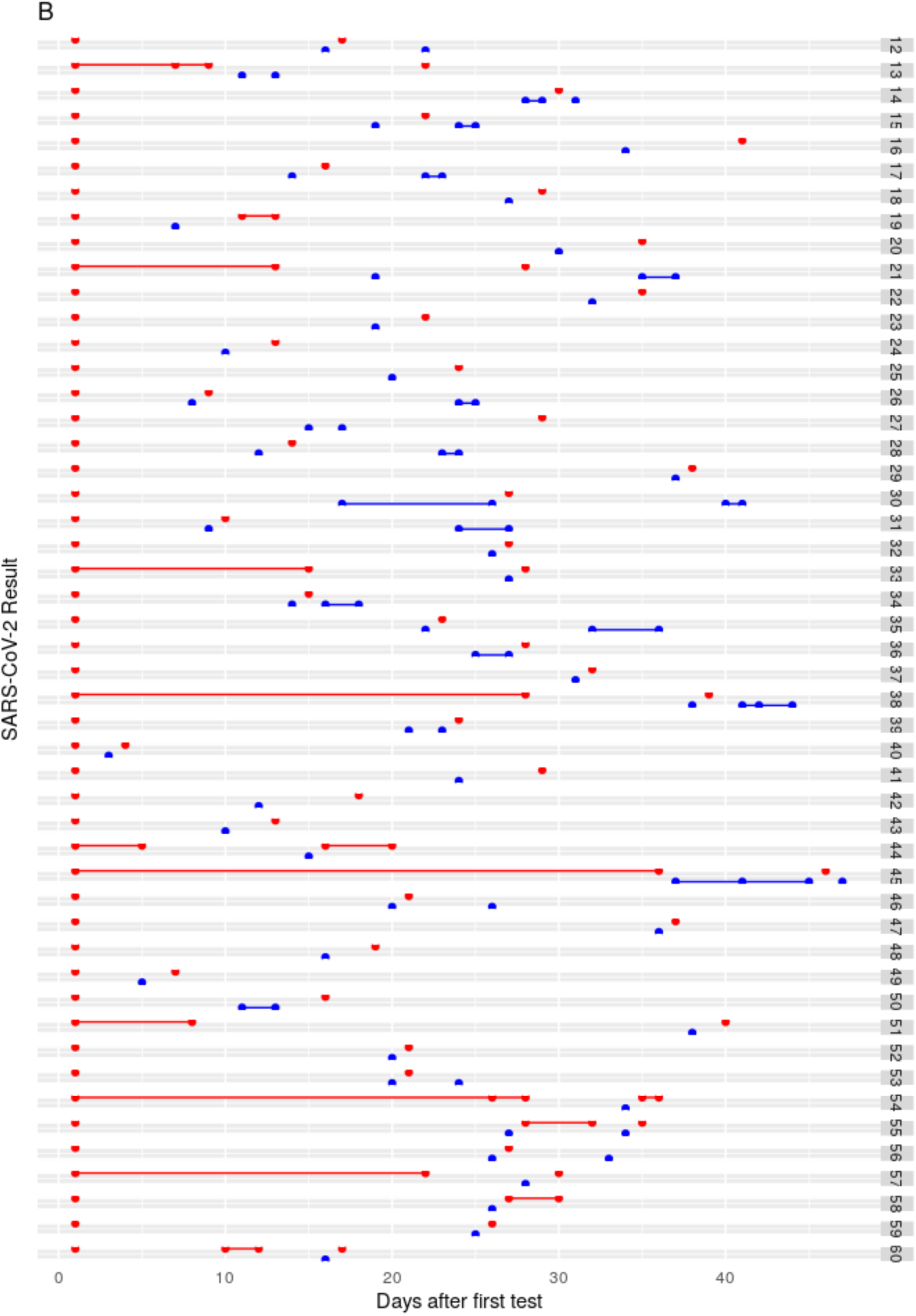
SARS-CoV-2 results from repeat-tested patients that converted from negative to positive and back to negative (**A**) or from positive to negative and back to positive (**B**). SARS-CoV-2 positive results are represented by blue dots and positive results are in red, plotted on a time scale from the date of the first test

## Supplementary Tables

**Supplementary Table 1:**
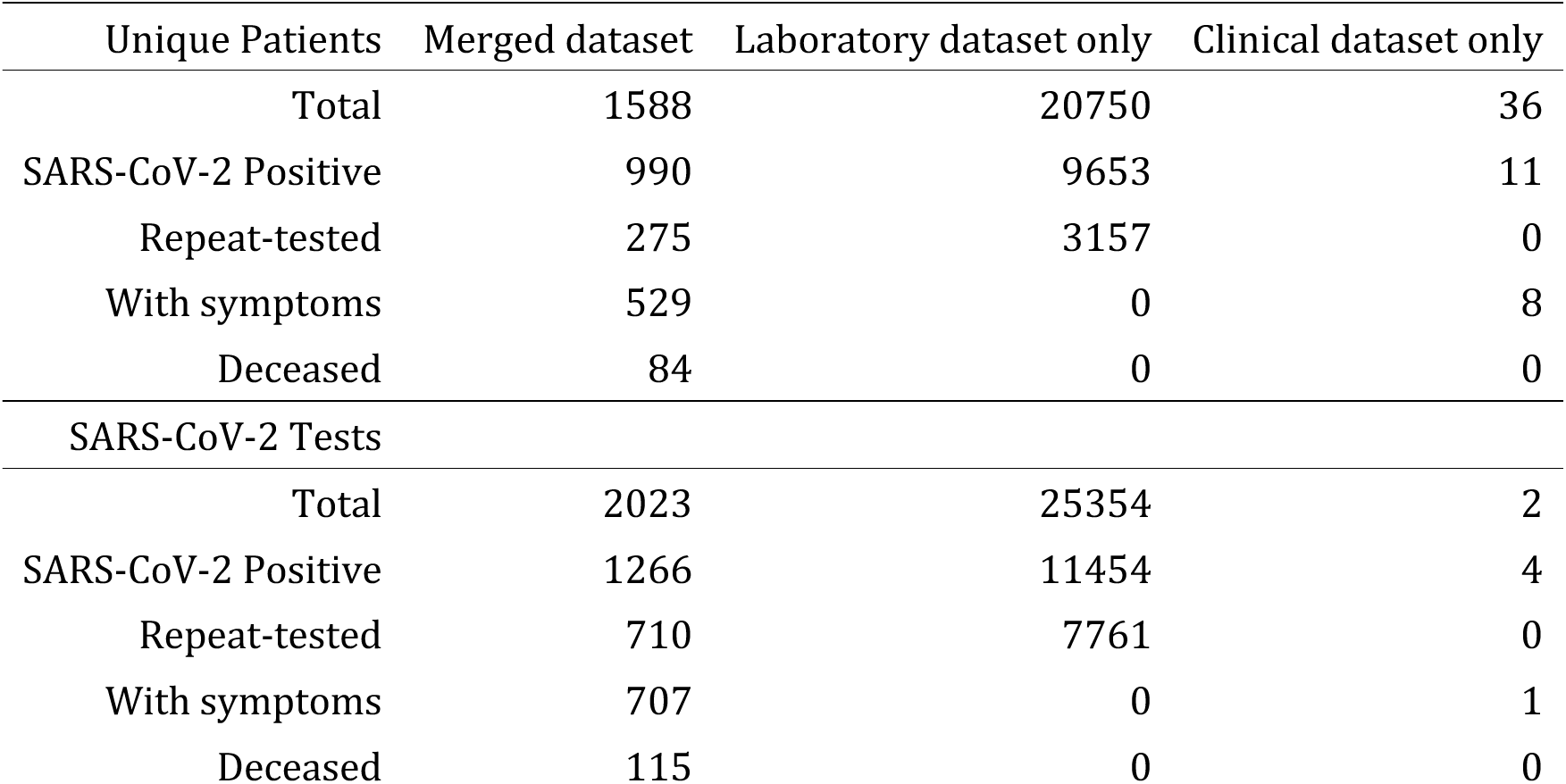
Numbers of unique patients in the Columbia Clinical COVID-19 dataset, NYP Laboratory SARS-CoV-2 Testing dataset, and the merged dataset.

| Unique Patients | Merged dataset | Laboratory dataset only | Clinical dataset only |
| --- | --- | --- | --- |
| Total | 1588 | 20750 | 36 |
| SARS-CoV-2 Positive | 990 | 9653 | 11 |
| Repeat-tested | 275 | 3157 | 0 |
| With symptoms | 529 | 0 | 8 |
| Deceased | 84 | 0 | 0 |
| SARS-CoV-2 Tests |  |  |  |
| Total | 2023 | 25354 | 2 |
| SARS-CoV-2 Positive | 1266 | 11454 | 4 |
| Repeat-tested | 710 | 7761 | 0 |
| With symptoms | 707 | 0 | 1 |
| Deceased | 115 | 0 | 0 |

**Supplementary Table 2:**
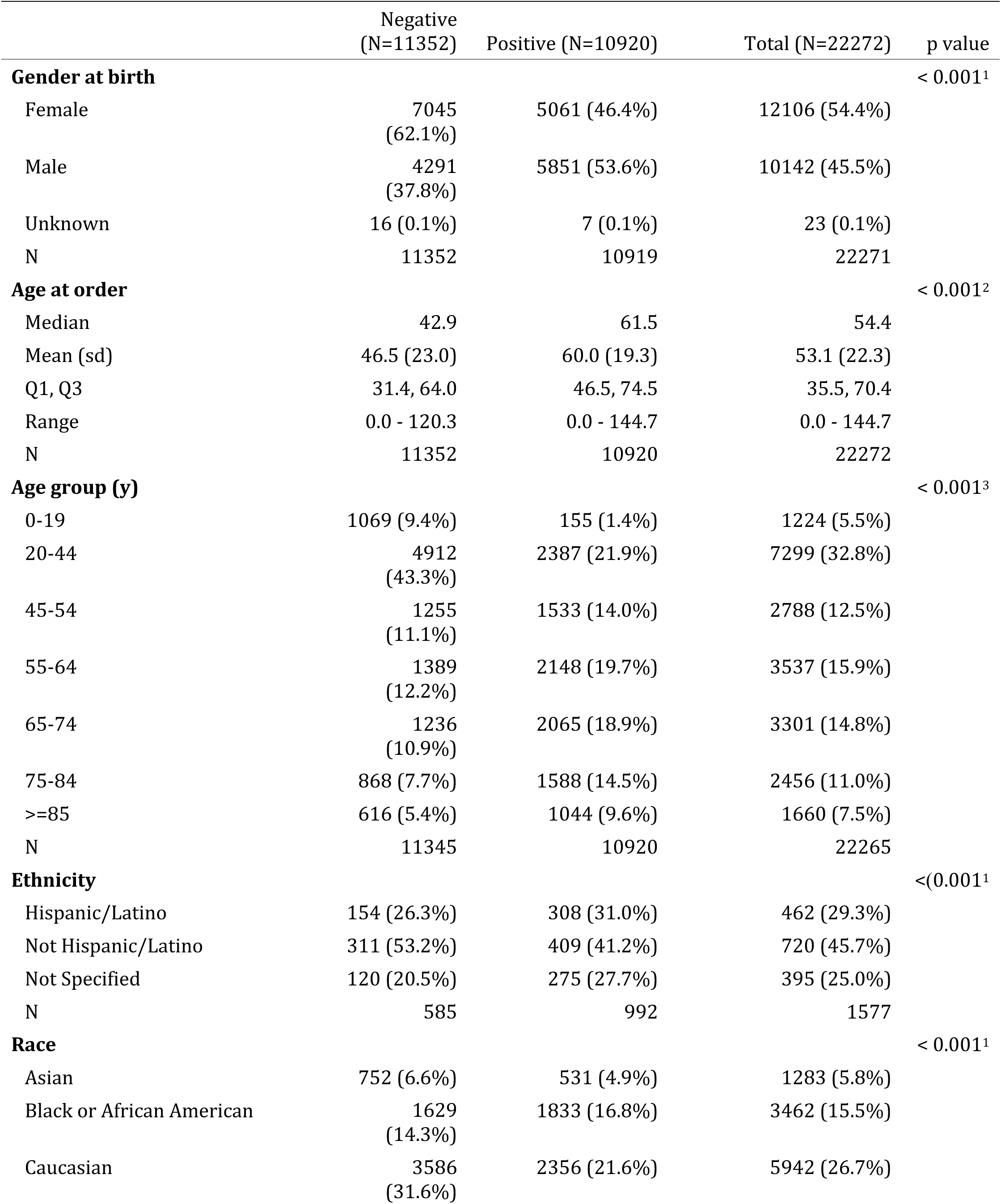
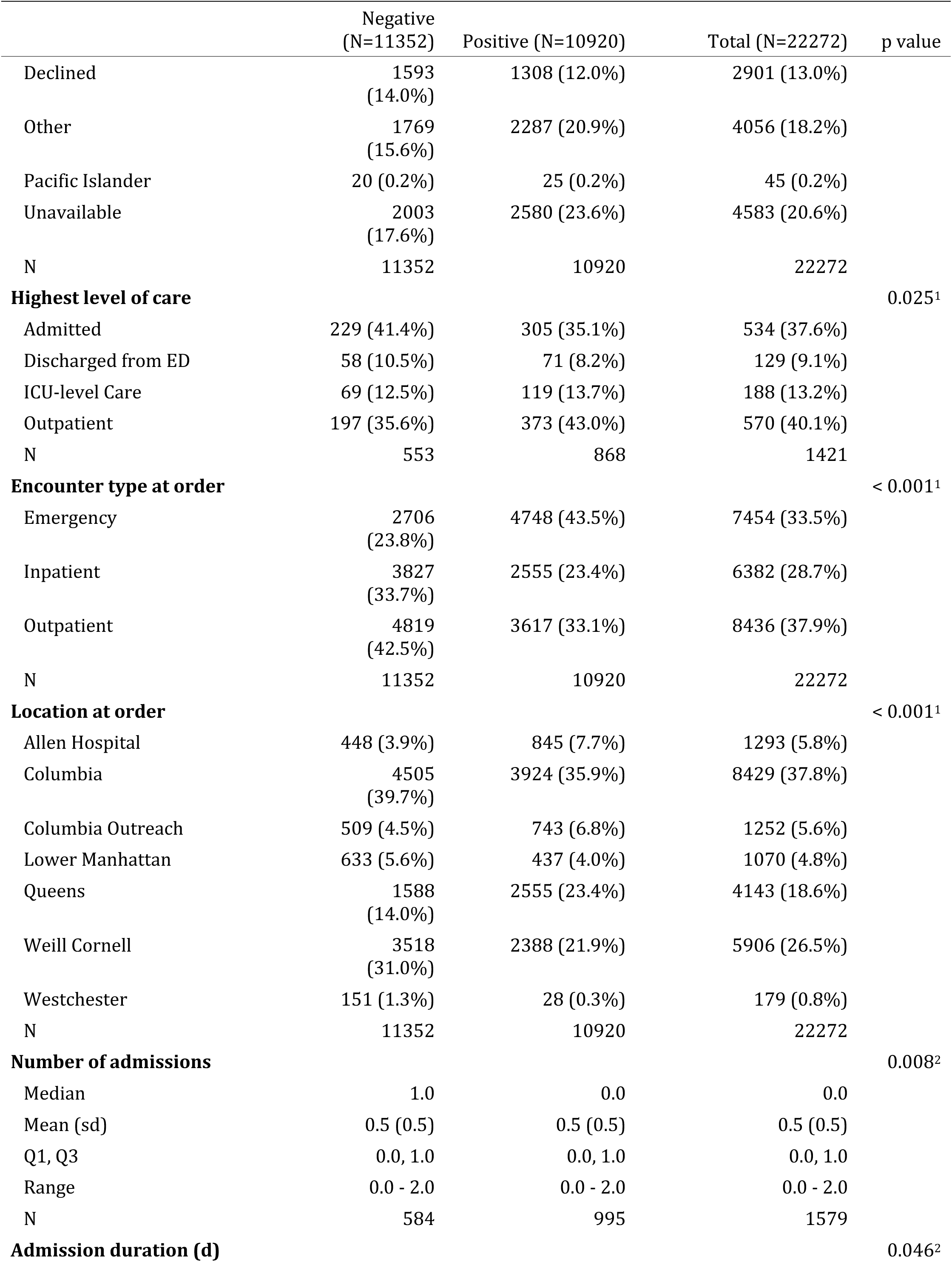
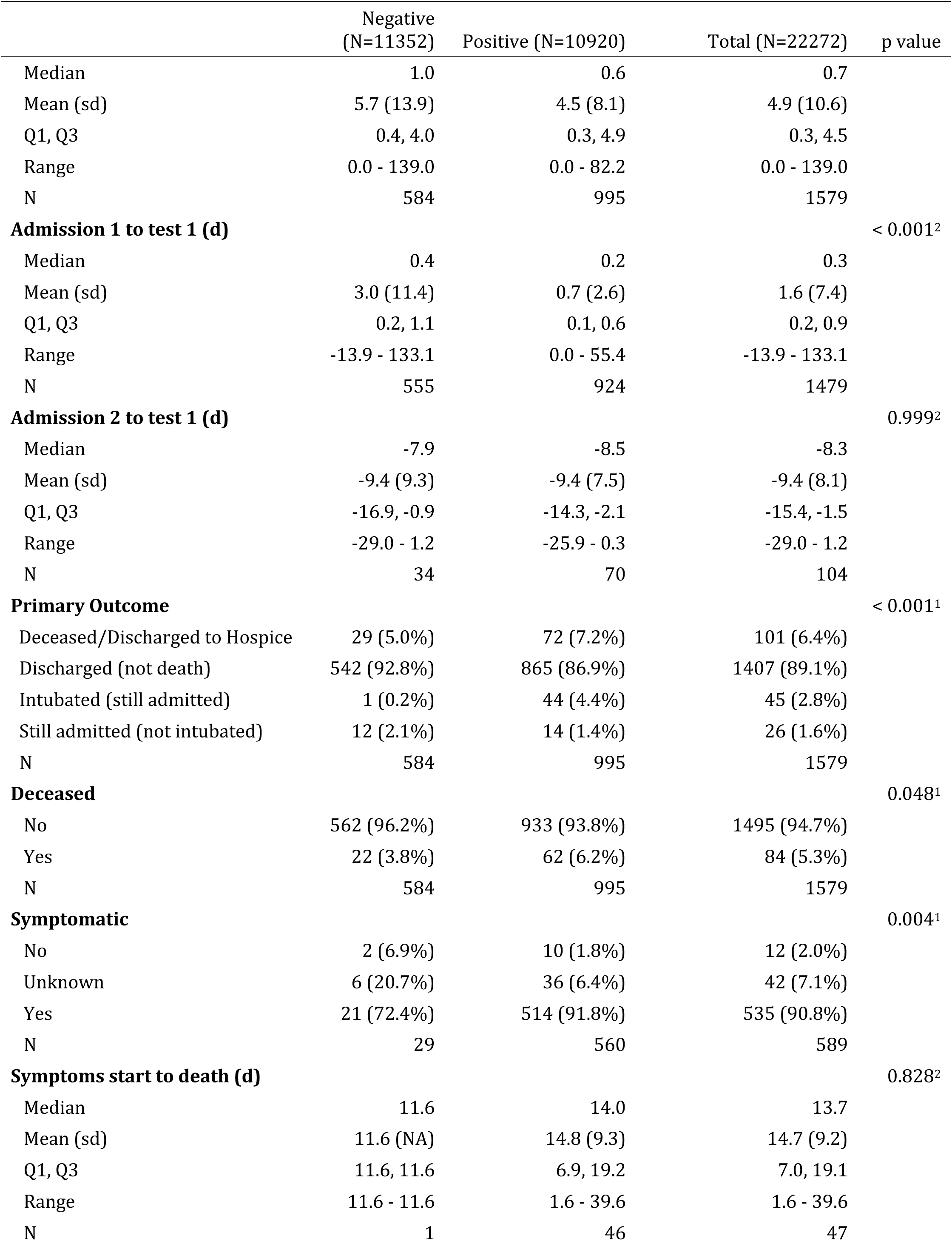
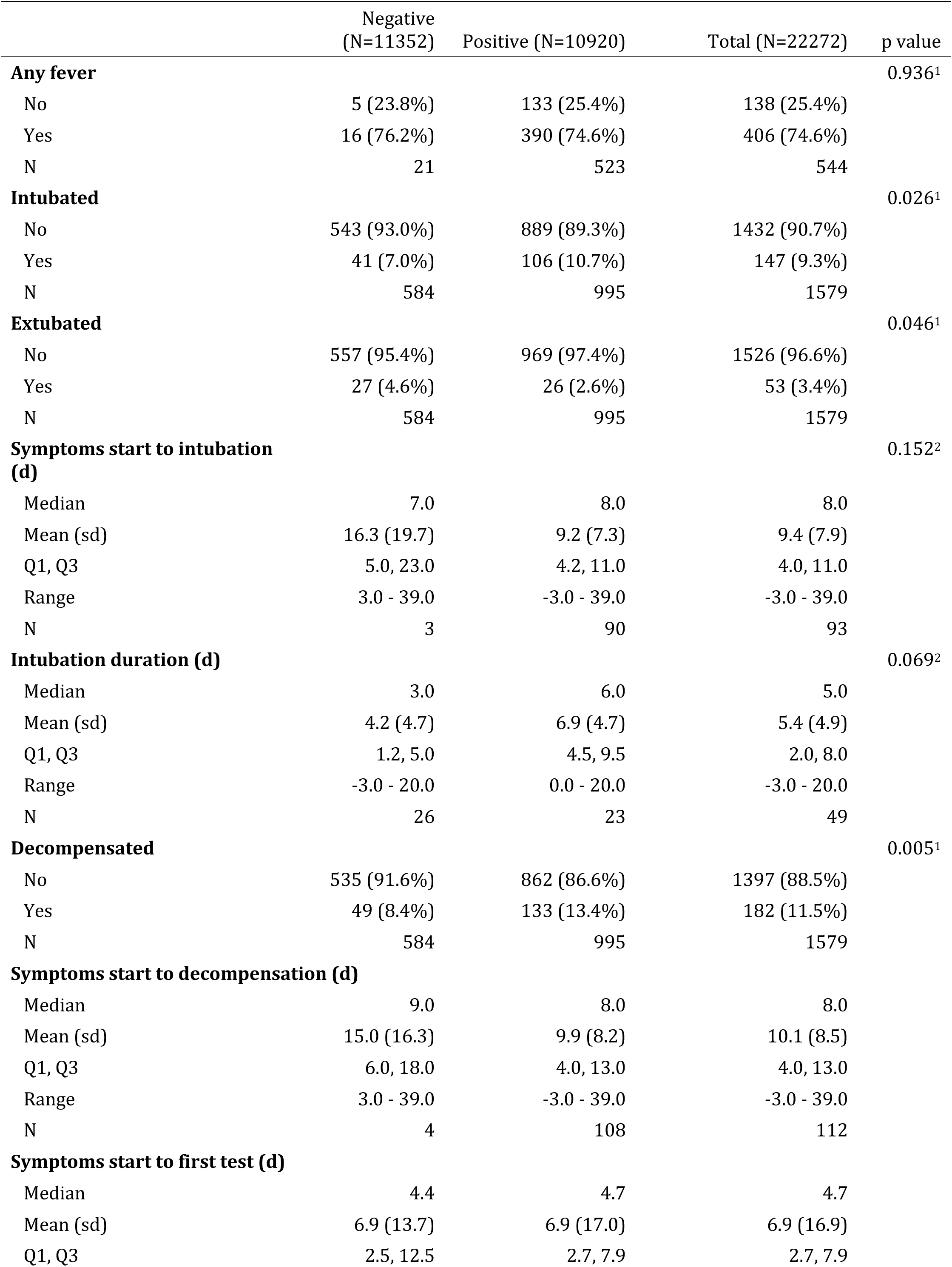
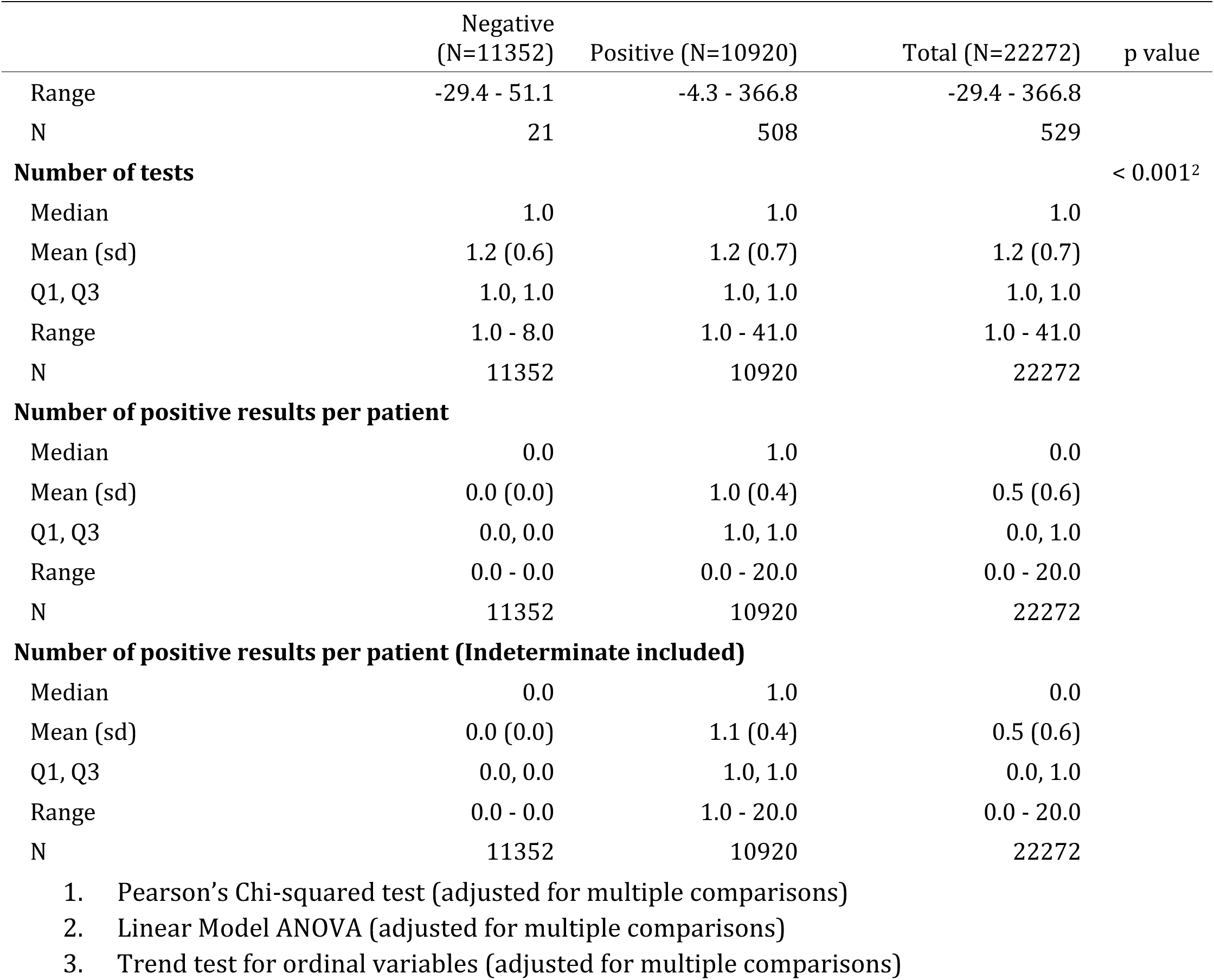
Patient characteristics, grouped by highest SARS-2-CoV2 result in each patient. **Negative** indicates a result of “Not Detected” and **Positive** includes results reported as “Detected” or “Indeterminate”. Time-dependent clinical factors, such as age group, symptom start to test interval, and encounter type at time of test order, are counted at the time of the first test.

|  | Negative<br>(N=11352) | Positive (N=10920) | Total (N=22272) | p value |
| --- | --- | --- | --- | --- |
| **Gender at birth** |  |  |  | < 0.001<sup>1</sup> |
| Female | 7045<br>(62.1%) | 5061 (46.4%) | 12106 (54.4%) |  |
| Male | 4291<br>(37.8%) | 5851 (53.6%) | 10142 (45.5%) |  |
| Unknown | 16 (0.1%) | 7 (0.1%) | 23 (0.1%) |  |
| N | 11352 | 10919 | 22271 |  |
| **Age at order** |  |  |  | < 0.001<sup>2</sup> |
| Median | 42.9 | 61.5 | 54.4 |  |
| Mean (sd) | 46.5 (23.0) | 60.0 (19.3) | 53.1 (22.3) |  |
| Q1, Q3 | 31.4, 64.0 | 46.5, 74.5 | 35.5, 70.4 |  |
| Range | 0.0 - 120.3 | 0.0 - 144.7 | 0.0 - 144.7 |  |
| N | 11352 | 10920 | 22272 |  |
| **Age group (y)** |  |  |  | < 0.001<sup>3</sup> |
| 0-19 | 1069 (9.4%) | 155 (1.4%) | 1224 (5.5%) |  |
| 20-44 | 4912<br>(43.3%) | 2387 (21.9%) | 7299 (32.8%) |  |
| 45-54 | 1255<br>(11.1%) | 1533 (14.0%) | 2788 (12.5%) |  |
| 55-64 | 1389<br>(12.2%) | 2148 (19.7%) | 3537 (15.9%) |  |
| 65-74 | 1236<br>(10.9%) | 2065 (18.9%) | 3301 (14.8%) |  |
| 75-84 | 868 (7.7%) | 1588 (14.5%) | 2456 (11.0%) |  |
| >=85 | 616 (5.4%) | 1044 (9.6%) | 1660 (7.5%) |  |
| N | 11345 | 10920 | 22265 |  |
| **Ethnicity** |  |  |  | <(0.001<sup>1</sup> |
| Hispanic/Latino | 154 (26.3%) | 308 (31.0%) | 462 (29.3%) |  |
| Not Hispanic/Latino | 311 (53.2%) | 409 (41.2%) | 720 (45.7%) |  |
| Not Specified | 120 (20.5%) | 275 (27.7%) | 395 (25.0%) |  |
| N | 585 | 992 | 1577 |  |
| **Race** |  |  |  | < 0.001<sup>1</sup> |
| Asian | 752 (6.6%) | 531 (4.9%) | 1283 (5.8%) |  |
| Black or African American | 1629<br>(14.3%) | 1833 (16.8%) | 3462 (15.5%) |  |
| Caucasian | 3586<br>(31.6%) | 2356 (21.6%) | 5942 (26.7%) |  |
| Declined | 1593<br>(14.0%) | 1308 (12.0%) | 2901 (13.0%) |  |
| Other | 1769<br>(15.6%) | 2287 (20.9%) | 4056 (18.2%) |  |
| Pacific Islander | 20 (0.2%) | 25 (0.2%) | 45 (0.2%) |  |
| Unavailable | 2003<br>(17.6%) | 2580 (23.6%) | 4583 (20.6%) |  |
| N | 11352 | 10920 | 22272 |  |
| **Highest level of care** |  |  |  | 0.025<sup>1</sup> |
| Admitted | 229 (41.4%) | 305 (35.1%) | 534 (37.6%) |  |
| Discharged from ED | 58 (10.5%) | 71 (8.2%) | 129 (9.1%) |  |
| ICU-level Care | 69 (12.5%) | 119 (13.7%) | 188 (13.2%) |  |
| Outpatient | 197 (35.6%) | 373 (43.0%) | 570 (40.1%) |  |
| N | 553 | 868 | 1421 |  |
| **Encounter type at order** |  |  |  | < 0.001<sup>1</sup> |
| Emergency | 2706<br>(23.8%) | 4748 (43.5%) | 7454 (33.5%) |  |
| Inpatient | 3827<br>(33.7%) | 2555 (23.4%) | 6382 (28.7%) |  |
| Outpatient | 4819<br>(42.5%) | 3617 (33.1%) | 8436 (37.9%) |  |
| N | 11352 | 10920 | 22272 |  |
| **Location at order** |  |  |  | < 0.001<sup>1</sup> |
| Allen Hospital | 448 (3.9%) | 845 (7.7%) | 1293 (5.8%) |  |
| Columbia | 4505<br>(39.7%) | 3924 (35.9%) | 8429 (37.8%) |  |
| Columbia Outreach | 509 (4.5%) | 743 (6.8%) | 1252 (5.6%) |  |
| Lower Manhattan | 633 (5.6%) | 437 (4.0%) | 1070 (4.8%) |  |
| Queens | 1588<br>(14.0%) | 2555 (23.4%) | 4143 (18.6%) |  |
| Weill Cornell | 3518<br>(31.0%) | 2388 (21.9%) | 5906 (26.5%) |  |
| Westchester | 151 (1.3%) | 28 (0.3%) | 179 (0.8%) |  |
| N | 11352 | 10920 | 22272 |  |
| **Number of admissions** |  |  |  | 0.008<sup>2</sup> |
| Median | 1.0 | 0.0 | 0.0 |  |
| Mean (sd) | 0.5 (0.5) | 0.5 (0.5) | 0.5 (0.5) |  |
| Q1, Q3 | 0.0, 1.0 | 0.0, 1.0 | 0.0, 1.0 |  |
| Range | 0.0 - 2.0 | 0.0 - 2.0 | 0.0 - 2.0 |  |
| N | 584 | 995 | 1579 |  |
| **Admission duration (d)** |  |  |  | 0.046<sup>2</sup> |
| Median | 1.0 | 0.6 | 0.7 |  |
| Mean (sd) | 5.7 (13.9) | 4.5 (8.1) | 4.9 (10.6) |  |
| Q1, Q3 | 0.4, 4.0 | 0.3, 4.9 | 0.3, 4.5 |  |
| Range | 0.0 - 139.0 | 0.0 - 82.2 | 0.0 - 139.0 |  |
| N | 584 | 995 | 1579 |  |
| **Admission 1 to test 1 (d)** |  |  |  | < 0.001<sup>2</sup> |
| Median | 0.4 | 0.2 | 0.3 |  |
| Mean (sd) | 3.0 (11.4) | 0.7 (2.6) | 1.6 (7.4) |  |
| Q1, Q3 | 0.2, 1.1 | 0.1, 0.6 | 0.2, 0.9 |  |
| Range | -13.9 - 133.1 | 0.0 - 55.4 | -13.9 - 133.1 |  |
| N | 555 | 924 | 1479 |  |
| **Admission 2 to test 1 (d)** |  |  |  | 0.999<sup>2</sup> |
| Median | -7.9 | -8.5 | -8.3 |  |
| Mean (sd) | -9.4 (9.3) | -9.4 (7.5) | -9.4 (8.1) |  |
| Q1, Q3 | -16.9, -0.9 | -14.3, -2.1 | -15.4, -1.5 |  |
| Range | -29.0 - 1.2 | -25.9 - 0.3 | -29.0 - 1.2 |  |
| N | 34 | 70 | 104 |  |
| **Primary Outcome** |  |  |  | < 0.001<sup>1</sup> |
| Deceased/Discharged to Hospice | 29 (5.0%) | 72 (7.2%) | 101 (6.4%) |  |
| Discharged (not death) | 542 (92.8%) | 865 (86.9%) | 1407 (89.1%) |  |
| Intubated (still admitted) | 1 (0.2%) | 44 (4.4%) | 45 (2.8%) |  |
| Still admitted (not intubated) | 12 (2.1%) | 14 (1.4%) | 26 (1.6%) |  |
| N | 584 | 995 | 1579 |  |
| **Deceased** |  |  |  | 0.048<sup>1</sup> |
| No | 562 (96.2%) | 933 (93.8%) | 1495 (94.7%) |  |
| Yes | 22 (3.8%) | 62 (6.2%) | 84 (5.3%) |  |
| N | 584 | 995 | 1579 |  |
| **Symptomatic** |  |  |  | 0.004<sup>1</sup> |
| No | 2 (6.9%) | 10 (1.8%) | 12 (2.0%) |  |
| Unknown | 6 (20.7%) | 36 (6.4%) | 42 (7.1%) |  |
| Yes | 21 (72.4%) | 514 (91.8%) | 535 (90.8%) |  |
| N | 29 | 560 | 589 |  |
| **Symptoms start to death (d)** |  |  |  | 0.828<sup>2</sup> |
| Median | 11.6 | 14.0 | 13.7 |  |
| Mean (sd) | 11.6 (NA) | 14.8 (9.3) | 14.7 (9.2) |  |
| Q1, Q3 | 11.6, 11.6 | 6.9, 19.2 | 7.0, 19.1 |  |
| Range | 11.6 - 11.6 | 1.6 - 39.6 | 1.6 - 39.6 |  |
| N | 1 | 46 | 47 |  |
| **Any fever** |  |  |  | 0.936<sup>1</sup> |
| No | 5 (23.8%) | 133 (25.4%) | 138 (25.4%) |  |
| Yes | 16 (76.2%) | 390 (74.6%) | 406 (74.6%) |  |
| N | 21 | 523 | 544 |  |
| **Intubated** |  |  |  | 0.026<sup>1</sup> |
| No | 543 (93.0%) | 889 (89.3%) | 1432 (90.7%) |  |
| Yes | 41 (7.0%) | 106 (10.7%) | 147 (9.3%) |  |
| N | 584 | 995 | 1579 |  |
| **Extubated** |  |  |  | 0.046<sup>1</sup> |
| No | 557 (95.4%) | 969 (97.4%) | 1526 (96.6%) |  |
| Yes | 27 (4.6%) | 26 (2.6%) | 53 (3.4%) |  |
| N | 584 | 995 | 1579 |  |
| **Symptoms start to intubation**<br>**(d)** |  |  |  | 0.152<sup>2</sup> |
| Median | 7.0 | 8.0 | 8.0 |  |
| Mean (sd) | 16.3 (19.7) | 9.2 (7.3) | 9.4 (7.9) |  |
| Q1, Q3 | 5.0, 23.0 | 4.2, 11.0 | 4.0, 11.0 |  |
| Range | 3.0 - 39.0 | -3.0 - 39.0 | -3.0 - 39.0 |  |
| N | 3 | 90 | 93 |  |
| **Intubation duration (d)** |  |  |  | 0.069<sup>2</sup> |
| Median | 3.0 | 6.0 | 5.0 |  |
| Mean (sd) | 4.2 (4.7) | 6.9 (4.7) | 5.4 (4.9) |  |
| Q1, Q3 | 1.2, 5.0 | 4.5, 9.5 | 2.0, 8.0 |  |
| Range | -3.0 - 20.0 | 0.0 - 20.0 | -3.0 - 20.0 |  |
| N | 26 | 23 | 49 |  |
| **Decompensated** |  |  |  | 0.005<sup>1</sup> |
| No | 535 (91.6%) | 862 (86.6%) | 1397 (88.5%) |  |
| Yes | 49 (8.4%) | 133 (13.4%) | 182 (11.5%) |  |
| N | 584 | 995 | 1579 |  |
| **Symptoms start to decompensation (d)** |  |  |  |  |
| Median | 9.0 | 8.0 | 8.0 |  |
| Mean (sd) | 15.0 (16.3) | 9.9 (8.2) | 10.1 (8.5) |  |
| Q1, Q3 | 6.0, 18.0 | 4.0, 13.0 | 4.0, 13.0 |  |
| Range | 3.0 - 39.0 | -3.0 - 39.0 | -3.0 - 39.0 |  |
| N | 4 | 108 | 112 |  |
| **Symptoms start to first test (d)** |  |  |  |  |
| Median | 4.4 | 4.7 | 4.7 |  |
| Mean (sd) | 6.9 (13.7) | 6.9 (17.0) | 6.9 (16.9) |  |
| Q1, Q3 | 2.5, 12.5 | 2.7, 7.9 | 2.7, 7.9 |  |
| Range | -29.4 - 51.1 | -4.3 - 366.8 | -29.4 - 366.8 |  |
| N | 21 | 508 | 529 |  |
| **Number of tests** |  |  |  | < 0.001<sup>2</sup> |
| Median | 1.0 | 1.0 | 1.0 |  |
| Mean (sd) | 1.2 (0.6) | 1.2 (0.7) | 1.2 (0.7) |  |
| Q1, Q3 | 1.0, 1.0 | 1.0, 1.0 | 1.0, 1.0 |  |
| Range | 1.0 - 8.0 | 1.0 - 41.0 | 1.0 - 41.0 |  |
| N | 11352 | 10920 | 22272 |  |
| **Number of positive results per patient** |  |  |  |  |
| Median | 0.0 | 1.0 | 0.0 |  |
| Mean (sd) | 0.0 (0.0) | 1.0 (0.4) | 0.5 (0.6) |  |
| Q1, Q3 | 0.0, 0.0 | 1.0, 1.0 | 0.0, 1.0 |  |
| Range | 0.0 - 0.0 | 0.0 - 20.0 | 0.0 - 20.0 |  |
| N | 11352 | 10920 | 22272 |  |
| **Number of positive results per patient (Indeterminate included)** |  |  |  |  |
| Median | 0.0 | 1.0 | 0.0 |  |
| Mean (sd) | 0.0 (0.0) | 1.1 (0.4) | 0.5 (0.6) |  |
| Q1, Q3 | 0.0, 0.0 | 1.0, 1.0 | 0.0, 1.0 |  |
| Range | 0.0 - 0.0 | 1.0 - 20.0 | 0.0 - 20.0 |  |
| N | 11352 | 10920 | 22272 |  |
| 1. | Pearson's Chi-squared test (adjusted for multiple comparisons) |  |  |  |
| 2. | Linear Model ANOVA (adjusted for multiple comparisons) |  |  |  |
| 3. | Trend test for ordinal variables (adjusted for multiple comparisons) |  |  |  |

**Supplementary Table 3:** Patient characteristics comparing repeat-tested with single-tested patients.

|  | Repeat (N=3432) | Single (N=18906) | Total (N=22338) | p value |
| --- | --- | --- | --- | --- |
| **Gender at birth** |  |  |  | <0.001<sup>1</sup> |
| Female | 1644 (47.9%) | 10503 (55.6%) | 12147 (54.4%) |  |
| Male | 1787 (52.1%) | 8380 (44.3%) | 10167 (45.5%) |  |
| Unknown | 1 (0.0%) | 22 (0.1%) | 23 (0.1%) |  |
| N | 3432 | 18905 | 22337 |  |
| **Age at order** |  |  |  | <0.001<sup>2</sup> |
| Median | 59.7 | 53.4 | 54.4 |  |
| Mean (sd) | 55.5 (23.3) | 52.7 (22.1) | 53.1 (22.3) |  |
| Q1, Q3 | 39.6, 72.3 | 35.0, 70.0 | 35.5, 70.4 |  |
| Range | 0.0 - 120.3 | 0.0 - 144.7 | 0.0 - 144.7 |  |
| N | 3432 | 18906 | 22338 |  |
| **Age group (y)** |  |  |  | <0.001<sup>3</sup> |
| 0-19 | 278 (8.1%) | 949 (5.0%) | 1227 (5.5%) |  |
| 20-44 | 751 (21.9%) | 6569 (34.8%) | 7320 (32.8%) |  |
| 45-54 | 413 (12.0%) | 2384 (12.6%) | 2797 (12.5%) |  |
| 55-64 | 604 (17.6%) | 2943 (15.6%) | 3547 (15.9%) |  |
| 65-74 | 673 (19.6%) | 2639 (14.0%) | 3312 (14.8%) |  |
| 75-84 | 450 (13.1%) | 2017 (10.7%) | 2467 (11.0%) |  |
| >=85 | 259 (7.6%) | 1402 (7.4%) | 1661 (7.4%) |  |
| N | 3428 | 18903 | 22331 |  |
| **Ethnicity** |  |  |  | 0.231<sup>1</sup> |
| Hispanic/Latino | 92 (33.5%) | 371 (28.3%) | 463 (29.2%) |  |
| Not Hispanic/Latino | 121 (44.0%) | 602 (46.0%) | 723 (45.6%) |  |
| Not Specified | 62 (22.5%) | 337 (25.7%) | 399 (25.2%) |  |
| N | 275 | 1310 | 1585 |  |
| **Race** |  |  |  | < 0.001<sup>1</sup> |
| Asian | 220 (6.4%) | 1065 (5.6%) | 1285 (5.8%) |  |
| Black or African American | 620 (18.1%) | 2856 (15.1%) | 3476 (15.6%) |  |
| Caucasian | 963 (28.1%) | 5006 (26.5%) | 5969 (26.7%) |  |
| Declined | 476 (13.9%) | 2432 (12.9%) | 2908 (13.0%) |  |
| Other | 633 (18.4%) | 3431 (18.1%) | 4064 (18.2%) |  |
| Pacific Islander | 12 (0.3%) | 34 (0.2%) | 46 (0.2%) |  |
| Unavailable | 508 (14.8%) | 4082 (21.6%) | 4590 (20.5%) |  |
| N | 3432 | 18906 | 22338 |  |
| **Highest level of care** |  |  |  | <0.001<sup>1</sup> |
| Admitted | 120 (45.1%) | 414 (35.6%) | 534 (37.4%) |  |
| Discharged from ED | 16 (6.0%) | 113 (9.7%) | 129 (9.0%) |  |
| ICU-level Care | 79 (29.7%) | 109 (9.4%) | 188 (13.2%) |  |
| Outpatient | 51 (19.2%) | 526 (45.3%) | 577 (40.4%) |  |
| N | 266 | 1162 | 1428 |  |
| **Encounter type at order** |  |  |  | <0.001<sup>1</sup> |
| Emergency | 1273 (37.1%) | 6197 (32.8%) | 7470 (33.4%) |  |
| Inpatient | 1400 (40.8%) | 4998 (26.4%) | 6398 (28.6%) |  |
| Outpatient | 759 (22.1%) | 7711 (40.8%) | 8470 (37.9%) |  |
| N | 3432 | 18906 | 22338 |  |
| **Location at order** |  |  |  | <0.001<sup>1</sup> |
| Allen Hospital | 204 (5.9%) | 1099 (5.8%) | 1303 (5.8%) |  |
| Columbia | 1434 (41.8%) | 7031 (37.2%) | 8465 (37.9%) |  |
| Columbia Outreach | 116 (3.4%) | 1144 (6.1%) | 1260 (5.6%) |  |
| Lower Manhattan | 161 (4.7%) | 911 (4.8%) | 1072 (4.8%) |  |
| NYHQ | 495 (14.4%) | 3654 (19.3%) | 4149 (18.6%) |  |
| Weill Cornell | 973 (28.4%) | 4936 (26.1%) | 5909 (26.5%) |  |
| WESTCHESTER | 49 (1.4%) | 131 (0.7%) | 180 (0.8%) |  |
| N | 3432 | 18906 | 22338 |  |
| **Number of admissions** |  |  |  | <0.001<sup>2</sup> |
| Median | 1.0 | 0.0 | 0.0 |  |
| Mean (sd) | 0.9 (0.6) | 0.4 (0.5) | 0.5 (0.5) |  |
| Q1, Q3 | 0.0, 1.0 | 0.0, 1.0 | 0.0, 1.0 |  |
| Range | 0.0 - 2.0 | 0.0 - 1.0 | 0.0 - 2.0 |  |
| N | 275 | 1312 | 1587 |  |
| **Admission duration (d)** |  |  |  | <0.001<sup>2</sup> |
| Median | 5.8 | 0.6 | 0.7 |  |
| Mean (sd) | 11.6 (13.5) | 3.5 (9.3) | 4.9 (10.6) |  |
| Q1, Q3 | 1.0, 19.4 | 0.3, 2.9 | 0.3, 4.4 |  |
| Range | 0.0 - 83.5 | 0.0 - 139.0 | 0.0 - 139.0 |  |
| N | 275 | 1312 | 1587 |  |
| **Admission 1 to test 1 (d)** |  |  |  | 0.112<sup>2</sup> |
| Median | 0.4 | 0.3 | 0.3 |  |
| Mean (sd) | 2.3 (6.7) | 1.4 (7.5) | 1.6 (7.3) |  |
| Q1, Q3 | 0.2, 1.1 | 0.2, 0.9 | 0.2, 0.9 |  |
| Range | -13.9 - 56.1 | 0.0 - 133.1 | -13.9 - 133.1 |  |
| N | 268 | 1218 | 1486 |  |
| **Admission 2 to test 1 (d)** |  |  |  | <0.001<sup>2</sup> |
| Median | -9.0 | 0.1 | -8.3 |  |
| Mean (sd) | -10.6 (7.8) | -2.6 (5.9) | -9.4 (8.1) |  |
| Q1, Q3 | -15.8, -4.0 | -1.2, 0.2 | -15.4, -1.5 |  |
| Range | -29.0 - 0.5 | -19.1 - 1.2 | -29.0 - 1.2 |  |
| N | 88 | 16 | 104 |  |
| **Primary Outcome** |  |  |  | <0.001<sup>1</sup> |
| Deceased/Discharged to Hospice | 27 (9.8%) | 74 (5.6%) | 101 (6.4%) |  |
| Discharged (not death) | 196 (71.3%) | 1219 (92.9%) | 1415 (89.2%) |  |
| Intubated (still admitted) | 31 (11.3%) | 14 (1.1%) | 45 (2.8%) |  |
| Still admitted (not intubated) | 21 (7.6%) | 5 (0.4%) | 26 (1.6%) |  |
| N | 275 | 1312 | 1587 |  |
| **Deceased** |  |  |  | 0.038<sup>1</sup> |
| No | 253 (92.0%) | 1250 (95.3%) | 1503 (94.7%) |  |
| Yes | 22 (8.0%) | 62 (4.7%) | 84 (5.3%) |  |
| N | 275 | 1312 | 1587 |  |
| **Symptomatic** |  |  |  | 0.035<sup>1</sup> |
| No | 4 (3.8%) | 8 (1.6%) | 12 (2.0%) |  |
| Unknown | 2 (1.9%) | 42 (8.6%) | 44 (7.4%) |  |
| Yes | 99 (94.3%) | 436 (89.7%) | 535 (90.5%) |  |
| N | 105 | 486 | 591 |  |
| **Symptoms start to death (d)** |  |  |  | 0.320<sup>2</sup> |
| Median | 12.3 | 14.3 | 13.7 |  |
| Mean (sd) | 17.4 (11.9) | 14.0 (8.4) | 14.7 (9.2) |  |
| Q1, Q3 | 9.8, 25.6 | 6.9, 19.1 | 7.0, 19.1 |  |
| Range | 3.3 - 39.6 | 1.6 - 38.3 | 1.6 - 39.6 |  |
| N | 10 | 37 | 47 |  |
| **Any fever** |  |  |  | 0.925<sup>1</sup> |
| No | 25 (25.0%) | 113 (25.5%) | 138 (25.4%) |  |
| Yes | 75 (75.0%) | 331 (74.5%) | 406 (74.6%) |  |
| N | 100 | 444 | 544 |  |
| **Intubated** |  |  |  | <0.001<sup>1</sup> |
| No | 208 (75.6%) | 1232 (93.9%) | 1440 (90.7%) |  |
| Yes | 67 (24.4%) | 80 (6.1%) | 147 (9.3%) |  |
| N | 275 | 1312 | 1587 |  |
| **Extubated** |  |  |  | <0.001<sup>1</sup> |
| No | 251 (91.3%) | 1283 (97.8%) | 1534 (96.7%) |  |
| Yes | 24 (8.7%) | 29 (2.2%) | 53 (3.3%) |  |
| N | 275 | 1312 | 1587 |  |
| **Symptoms start to intubation (d)** |  |  |  |  |
| Median | 9.0 | 7.0 | 8.0 |  |
| Mean (sd) | 10.9 (7.0) | 8.3 (8.4) | 9.4 (7.9) |  |
| Q1, Q3 | 7.0, 14.0 | 3.0, 10.2 | 4.0, 11.0 |  |
| Range | 0.0 - 30.0 | -3.0 - 39.0 | -3.0 - 39.0 |  |
| N | 41 | 52 | 93 |  |
| **Intubation duration (d)** |  |  |  | 0.297<sup>2</sup> |
| Median | 5.0 | 4.5 | 5.0 |  |
| Mean (sd) | 6.3 (5.5) | 4.7 (4.2) | 5.4 (4.9) |  |
| Q1, Q3 | 2.5, 8.5 | 1.0, 7.8 | 2.0, 8.0 |  |
| Range | 1.0 - 20.0 | -3.0 - 13.0 | -3.0 - 20.0 |  |
| N | 23 | 26 | 49 |  |
| **Decompensated** |  |  |  | <0.001<sup>1</sup> |
| No | 199 (72.4%) | 1206 (91.9%) | 1405 (88.5%) |  |
| Yes | 76 (27.6%) | 106 (8.1%) | 182 (11.5%) |  |
| N | 275 | 1312 | 1587 |  |
| **Symptoms start to decompensation (d)** |  |  |  |  |
| Median | 9.0 | 7.0 | 8.0 |  |
| Mean (sd) | 11.7 (8.3) | 9.0 (8.6) | 10.1 (8.5) |  |
| Q1, Q3 | 7.0, 15.0 | 3.5, 11.0 | 4.0, 13.0 |  |
| Range | 0.0 - 39.0 | -3.0 - 39.0 | -3.0 - 39.0 |  |
| N | 45 | 67 | 112 |  |
| **Symptoms start to first test (d)** |  |  |  |  |
| Median | 5.3 | 4.6 | 4.7 |  |
| Mean (sd) | 9.8 (36.8) | 6.3 (6.4) | 6.9 (16.9) |  |
| Q1, Q3 | 2.1, 8.3 | 2.8, 7.8 | 2.7, 7.9 |  |
| Range | -4.3 - 366.8 | -29.4 - 60.8 | -29.4 - 366.8 |  |
| N | 98 | 431 | 529 |  |
| **Number of tests** |  |  |  | <0.001<sup>2</sup> |
| Median | 2.0 | 1.0 | 1.0 |  |
| Mean (sd) | 2.5 (1.1) | 1.0 (0.0) | 1.2 (0.7) |  |
| Q1, Q3 | 2.0, 3.0 | 1.0, 1.0 | 1.0, 1.0 |  |
| Range | 1.0 - 41.0 | 1.0 - 1.0 | 1.0 - 41.0 |  |
| N | 3432 | 18906 | 22338 |  |
| **Number of positive results per patient** |  |  |  |  |
| Median | 0.0 | 0.0 | 0.0 |  |
| Mean (sd) | 0.6 (0.9) | 0.5 (0.5) | 0.5 (0.6) |  |
| Q1, Q3 | 0.0, 1.0 | 0.0, 1.0 | 0.0, 1.0 |  |
| Range | 0.0 - 20.0 | 0.0 - 1.0 | 0.0 - 20.0 |  |
| N | 3432 | 18906 | 22338 |  |
| **Number of positive results per patient (indeterminate included)** |  |  |  |  |
| Median | 0.0 | 0.0 | 0.0 |  |
| Mean (sd) | 0.7 (1.0) | 0.5 (0.5) | 0.5 (0.6) |  |
| Q1, Q3 | 0.0, 1.0 | 0.0, 1.0 | 0.0, 1.0 |  |
| Range | 0.0 - 20.0 | 0.0 - 1.0 | 0.0 - 20.0 |  |
| N | 3432 | 18906 | 22338 | 1. |
| **Highest SARS-CoV-2 Result** |  |  |  | <0.001<sup>1</sup> |
| Invalid | 1 (0.0%) | 65 (0.3%) | 66 (0.3%) |  |
| Not Detected | 1960 (57.1%) | 9392 (49.7%) | 11352 (50.8%) |  |
| Indeterminate | 100 (2.9%) | 177 (0.9%) | 277 (1.2%) |  |
| Detected | 1371 (39.9%) | 9272 (49.0%) | 10643 (47.6%) |  |
| N | 3432 | 18906 | 22338 |  |
|  | Pearson's Chi-squared test (adjusted for multiple comparisons) |  |  |  |
| 2. | Linear Model ANOVA (adjusted for multiple comparisons) |  |  |  |
| 3. | Trend test for ordinal variables (adjusted for multiple comparisons) |  |  |  |

**Supplementary Table 4:** SARS-CoV-2 molecular tests in repeat-tested vs. single-tested patients. N represents the number of tests in each category.

|  | Repeat (N=8471) | Single (N=18906) | Total (N=27377) | p value |
| --- | --- | --- | --- | --- |
| **Method** |  |  |  | <0.001<sup>1</sup> |
| cobas 6800 | 5841 (69.0%) | 13354 (70.6%) | 19195 (70.1%) |  |
| Infiniti | 1900 (22.4%) | 4054 (21.4%) | 5954 (21.7%) |  |
| GeneXpert | 91 (1.1%) | 174 (0.9%) | 265 (1.0%) |  |
| ID Now | 9 (0.1%) | 44 (0.2%) | 53 (0.2%) |  |
| Panther Fusion | 11 (0.1%) | 15 (0.1%) | 26 (0.1%) |  |
| QuantStudio | 571 (6.7%) | 1224 (6.5%) | 1795 (6.6%) |  |
| 7500 Fast | 48 (0.6%) | 41 (0.2%) | 89 (0.3%) |  |
| N | 8471 | 18906 | 27377 |  |
| **cobas** |  |  |  | <0.001<sup>2</sup> |
| No | 8471 (100.0%) | 18906 (100.0%) | 27377 (100.0%) |  |
| N | 8471 | 18906 | 27377 |  |
| **Initial result** |  |  |  | <0.001<sup>1</sup> |
| Invalid | 554 (6.5%) | 65 (0.3%) | 619 (2.3%) |  |
| Not Detected | 5613 (66.3%) | 9392 (49.7%) | 15005 (54.8%) |  |
| Indeterminate | 265 (3.1%) | 177 (0.9%) | 442 (1.6%) |  |
| Detected | 2039 (24.1%) | 9272 (49.0%) | 11311 (41.3%) |  |
| N | 8471 | 18906 | 27377 |  |
| **Highest result on day 1** |  |  |  | <0.001<sup>1</sup> |
| Invalid | 367 (4.3%) | 65 (0.3%) | 432 (1.6%) |  |
| Not Detected | 5501 (64.9%) | 9392 (49.7%) | 14893 (54.4%) |  |
| Indeterminate | 267 (3.2%) | 177 (0.9%) | 444 (1.6%) |  |
| Detected | 2336 (27.6%) | 9272 (49.0%) | 11608 (42.4%) |  |
| N | 8471 | 18906 | 27377 |  |
| **Highest result any day** |  |  |  | <0.001<sup>1</sup> |
| Invalid | 2 (0.0%) | 65 (0.3%) | 67 (0.2%) |  |
| Not Detected | 4775 (56.4%) | 9392 (49.7%) | 14167 (51.7%) |  |
| Indeterminate | 246 (2.9%) | 177 (0.9%) | 423 (1.5%) |  |
| Detected | 3448 (40.7%) | 9272 (49.0%) | 12720 (46.5%) |  |
| N | 8471 | 18906 | 27377 |  |
| **Control Ct** |  |  |  | <0.001<sup>3</sup> |
| Median | 34.0 | 33.9 | 33.9 |  |
| Mean (sd) | 34.2 (1.1) | 34.1 (1.0) | 34.1 (1.0) |  |
| Q1, Q3 | 33.5, 34.7 | 33.4, 34.5 | 33.5, 34.6 |  |
| Range | 32.5 - 41.2 | 32.4 - 40.5 | 32.4 - 41.2 |  |
| N | 2144 | 7262 | 9406 |  |
| **Target 1 Ct** |  |  |  | 0.036<sup>3</sup> |
| Median | 27.5 | 26.6 | 26.7 |  |
| Mean (sd) | 26.7 (5.5) | 26.2 (5.5) | 26.3 (5.5) |  |
| Q1, Q3 | 22.3, 31.4 | 21.8, 31.0 | 21.9, 31.0 |  |
| Range | 13.3 - 36.8 | 12.4 - 38.0 | 12.4 - 38.0 |  |
| N | 744 | 4533 | 5277 |  |
| **Target 2 Ct** |  |  |  | <0.001<sup>3</sup> |
| Median | 29.1 | 27.3 | 27.6 |  |
| Mean (sd) | 28.5 (6.5) | 27.3 (6.1) | 27.5 (6.2) |  |
| Q1, Q3 | 23.2, 34.0 | 22.3, 32.5 | 22.5, 32.7 |  |
| Range | 13.7 - 39.8 | 12.9 - 43.7 | 12.9 - 43.7 |  |
| N | 795 | 4548 | 5343 |  |
| **Target 2 Ct > 30** |  |  |  | <0.001<sup>1</sup> |
| FALSE | 431 (54.2%) | 2869 (63.1%) | 3300 (61.8%) |  |
| TRUE | 364 (45.8%) | 1679 (36.9%) | 2043 (38.2%) |  |
| N | 795 | 4548 | 5343 |  |
| 1. | Pearson's Chi-squared test (adjusted for multiple comparisons) |  |  |  |
| 2. | Chi-squared test for given probabilities (adjusted for multiple comparisons) |  |  |  |
| 3. | Linear Model ANOVA (adjusted for multiple comparisons) |  |  |  |

**Supplementary Table 5:**
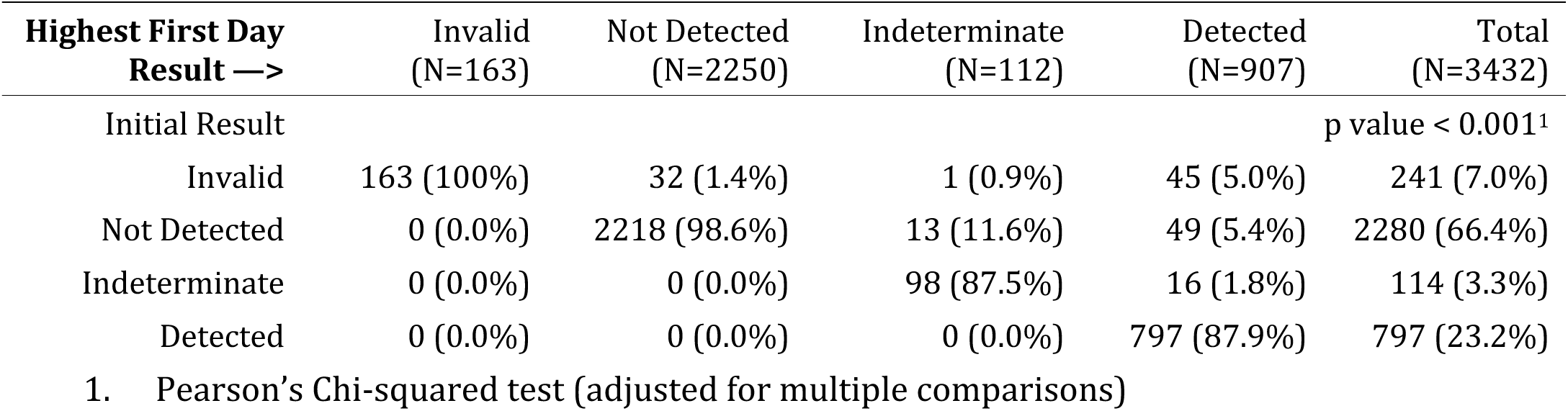
Number of SARS-CoV-2 molecular test results of repeat-tested patients on day 1 of testing. Rows represent the initial test result and columns the highest test result on day 1.

**Supplementary Table 6:** Number of SARS-CoV-2 molecular test results over the course of repeat testing, grouped by the initial test result on day 1.

| **Highest Result Any Day —>** | Invalid<br>(N=1) | Not Detected<br>(N=1960) | Indeterminate<br>(N=100) | Detected<br>(N=1371) | Total (N=3432) |
| --- | --- | --- | --- | --- | --- |
| Initial Result | p-value < 0.001<sup>1</sup> |  |  |  |  |
| Invalid | 1 (100%) | 110 (5.6%) | 3 (3.0%) | 127 (9.3%) | 241 (7.0%) |
| Not Detected | 0 (0.0%) | 1850 (94.4%) | 45 (45.0%) | 385 (28.1%) | 2280 (66.4%) |
| Indeterminate | 0 (0.0%) | 0 (0.0%) | 52 (52.0%) | 62 (4.5%) | 114 (3.3%) |
| Detected | 0 (0.0%) | 0 (0.0%) | 0 (0.0%) | 797 (58.1%) | 797 (23.2%) |
| 1. Pearson's Chi-squared test (adjusted for multiple comparisons) |  |  |  |  |  |

**Supplementary Table 7:**
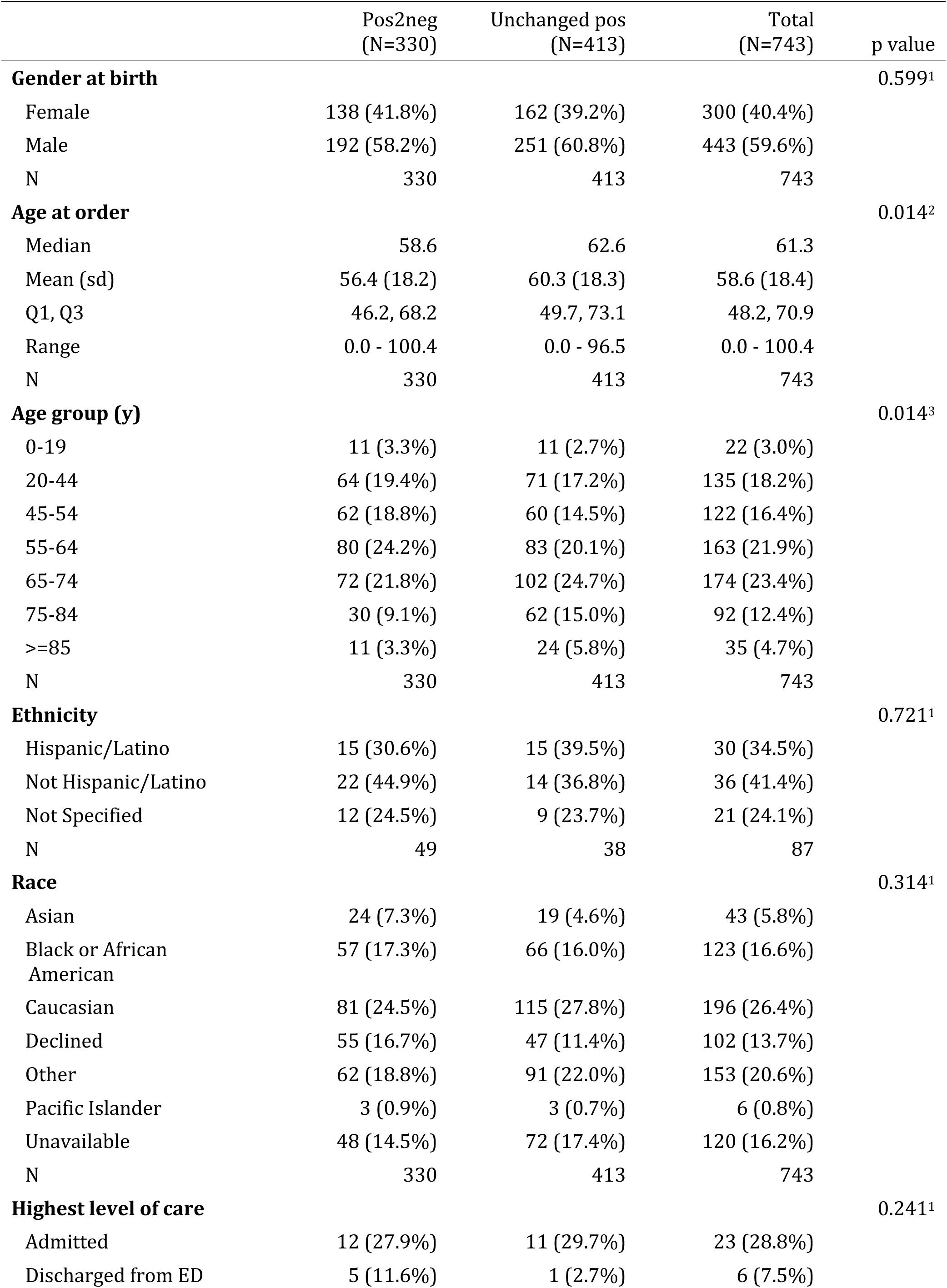
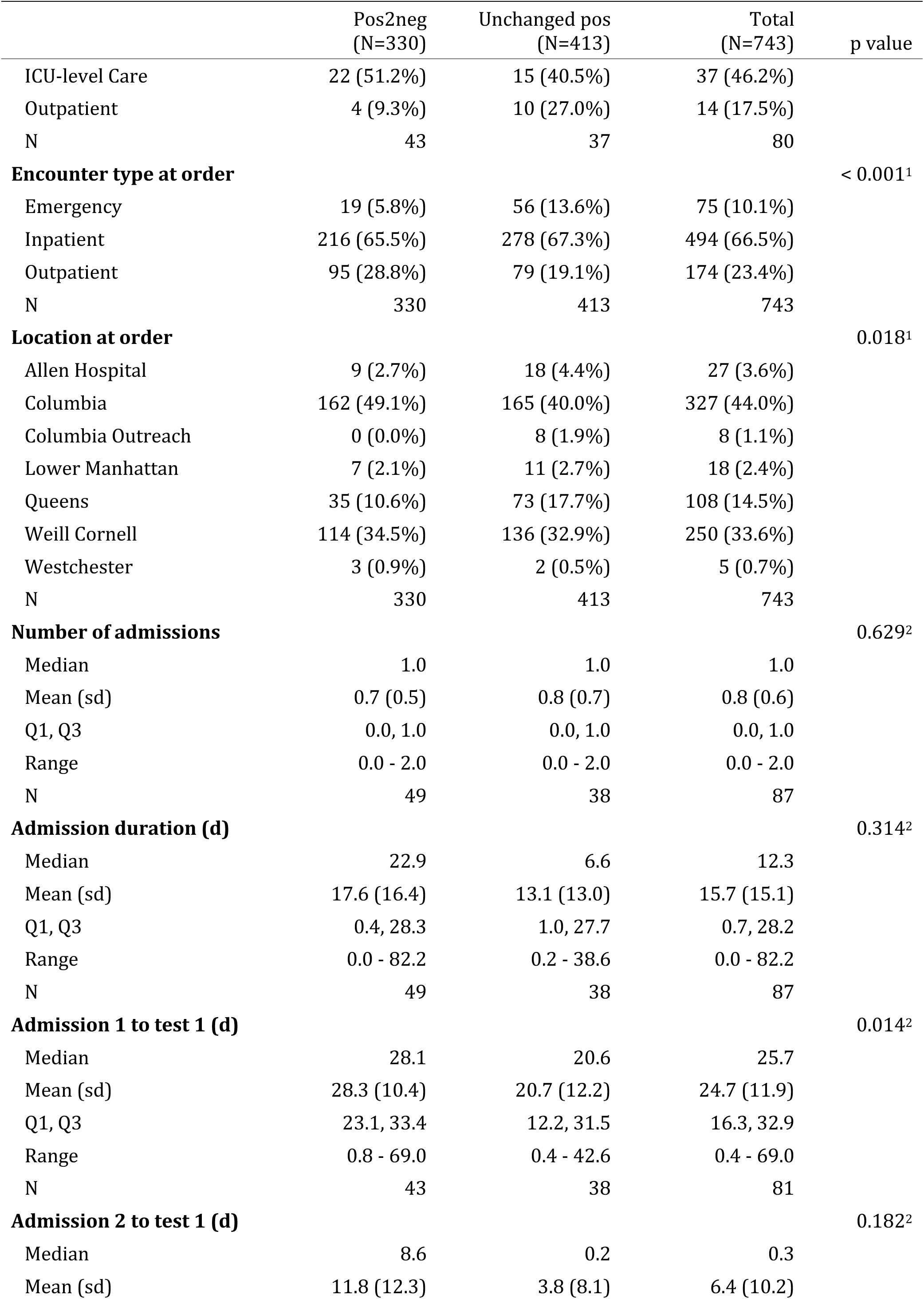
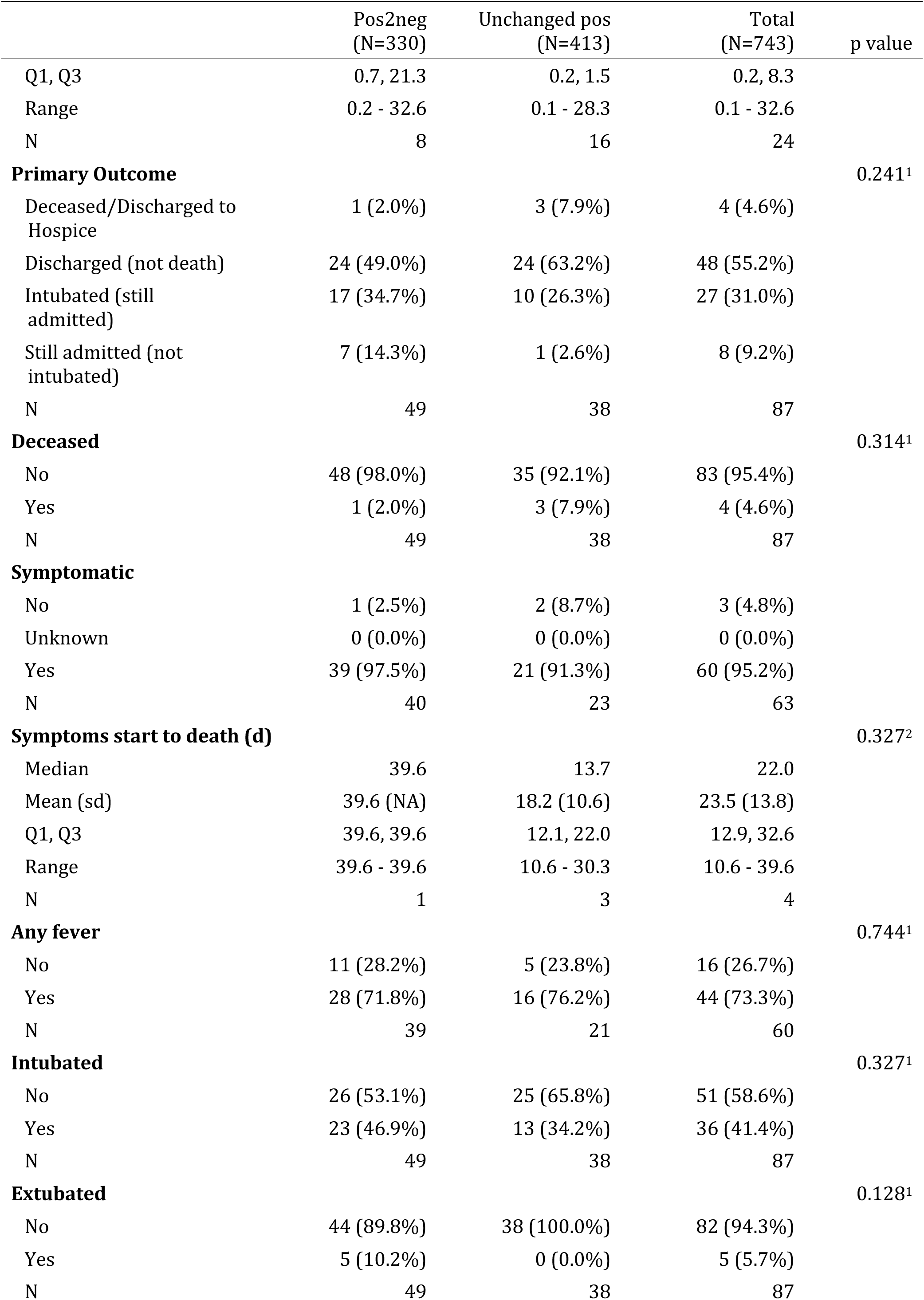
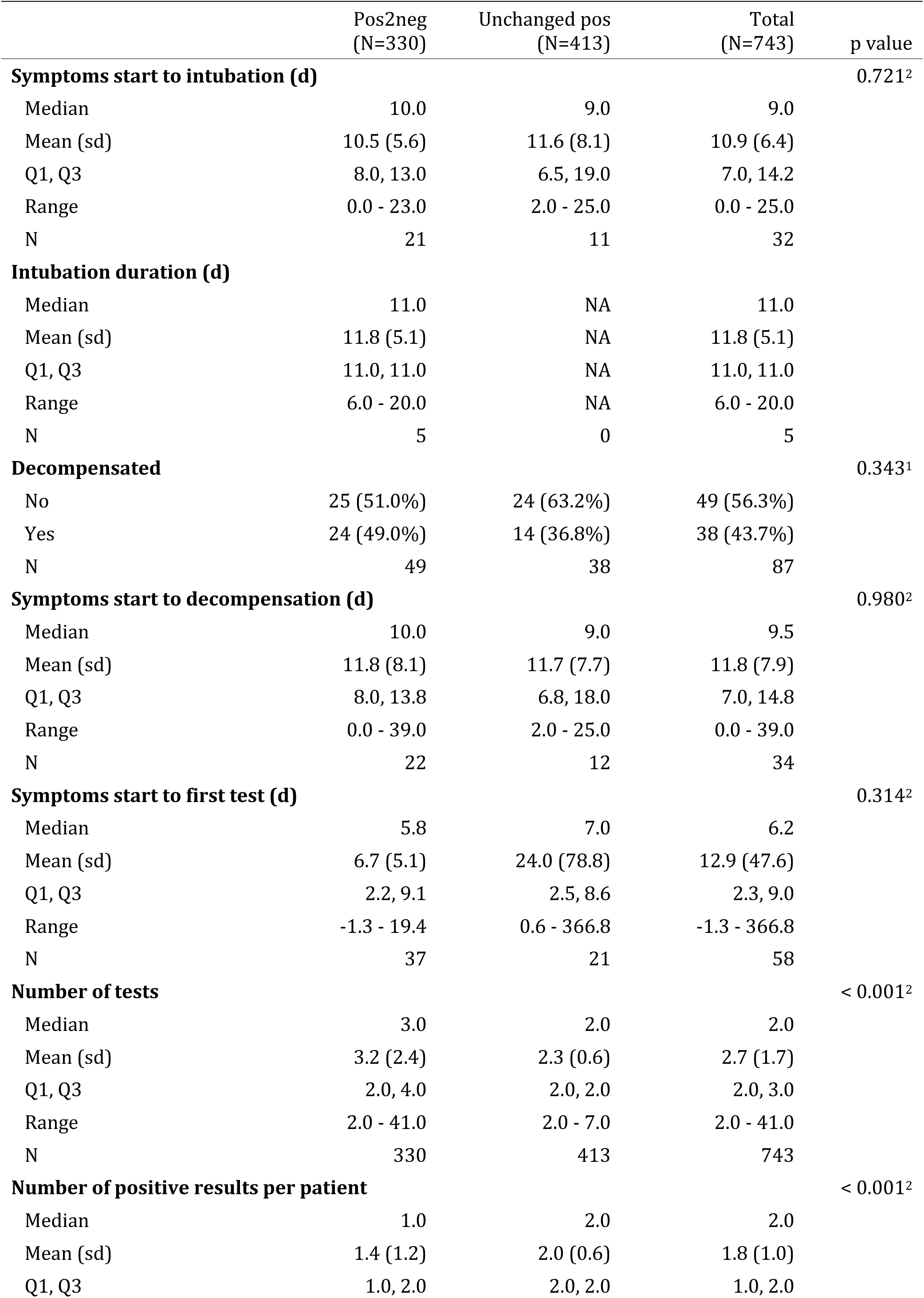
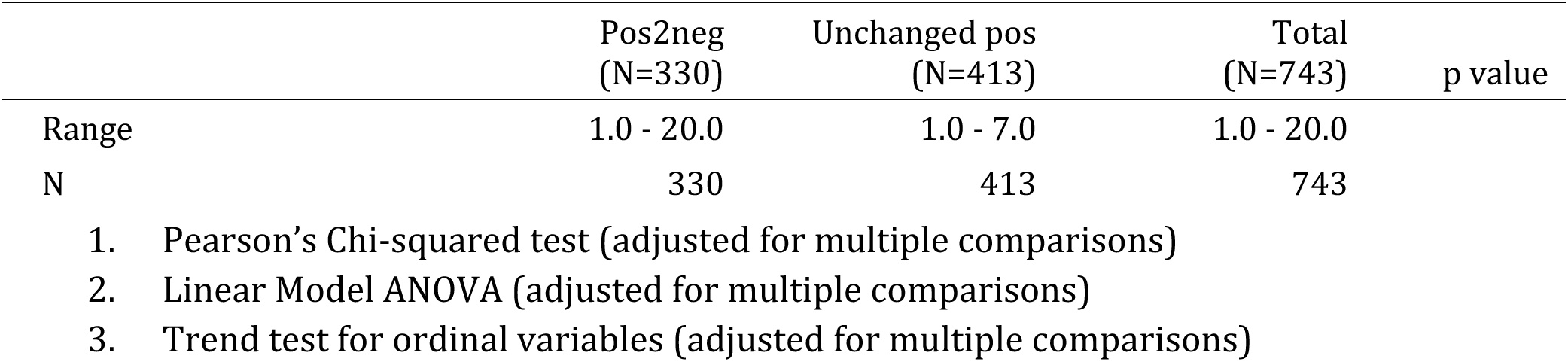
Characteristics of initially positive repeat-tested patients comparing those that converted from positive to negative (Pos2neg) with those that remained positive (Unchanged pos).

**Supplementary Table 8:**
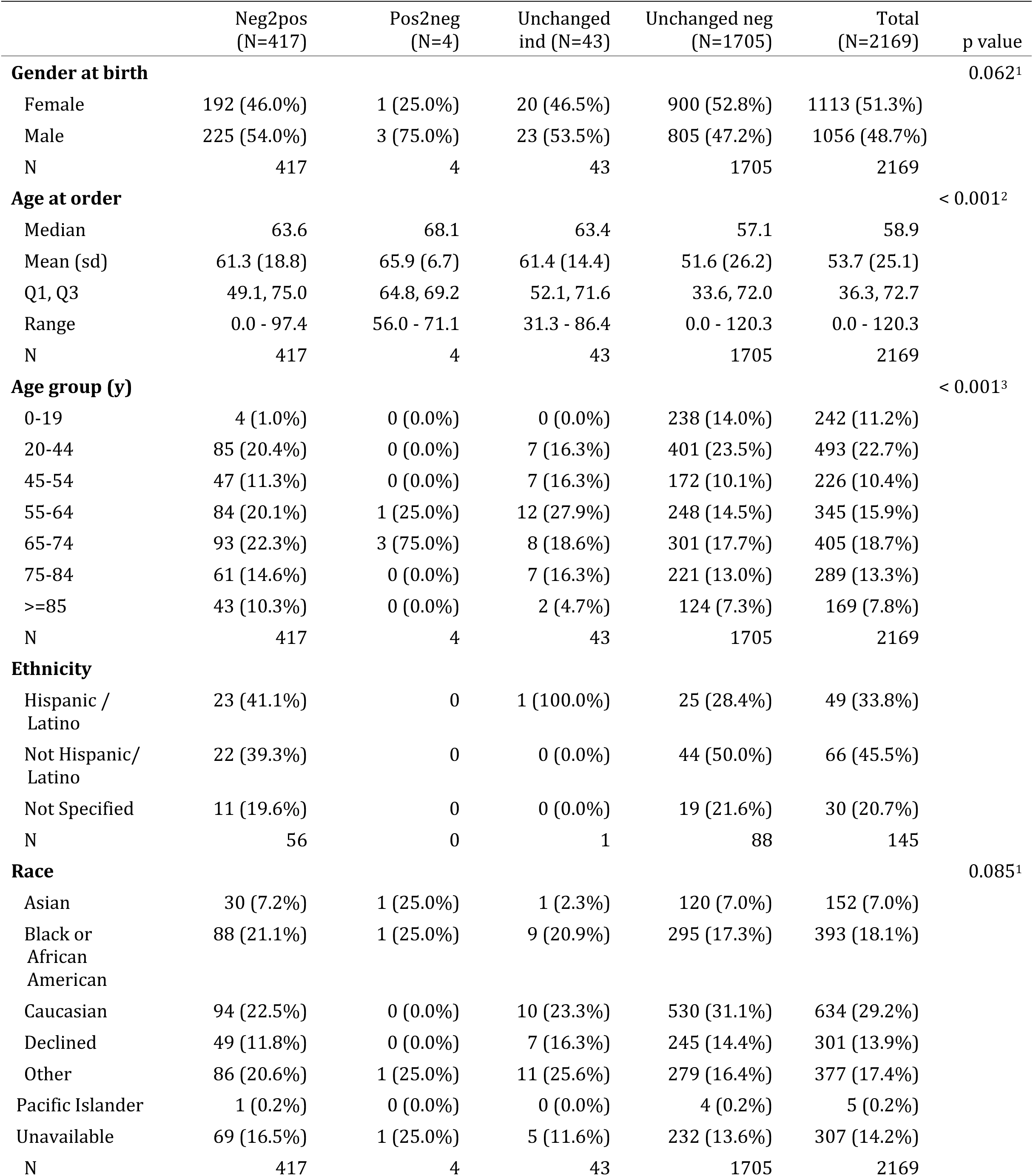
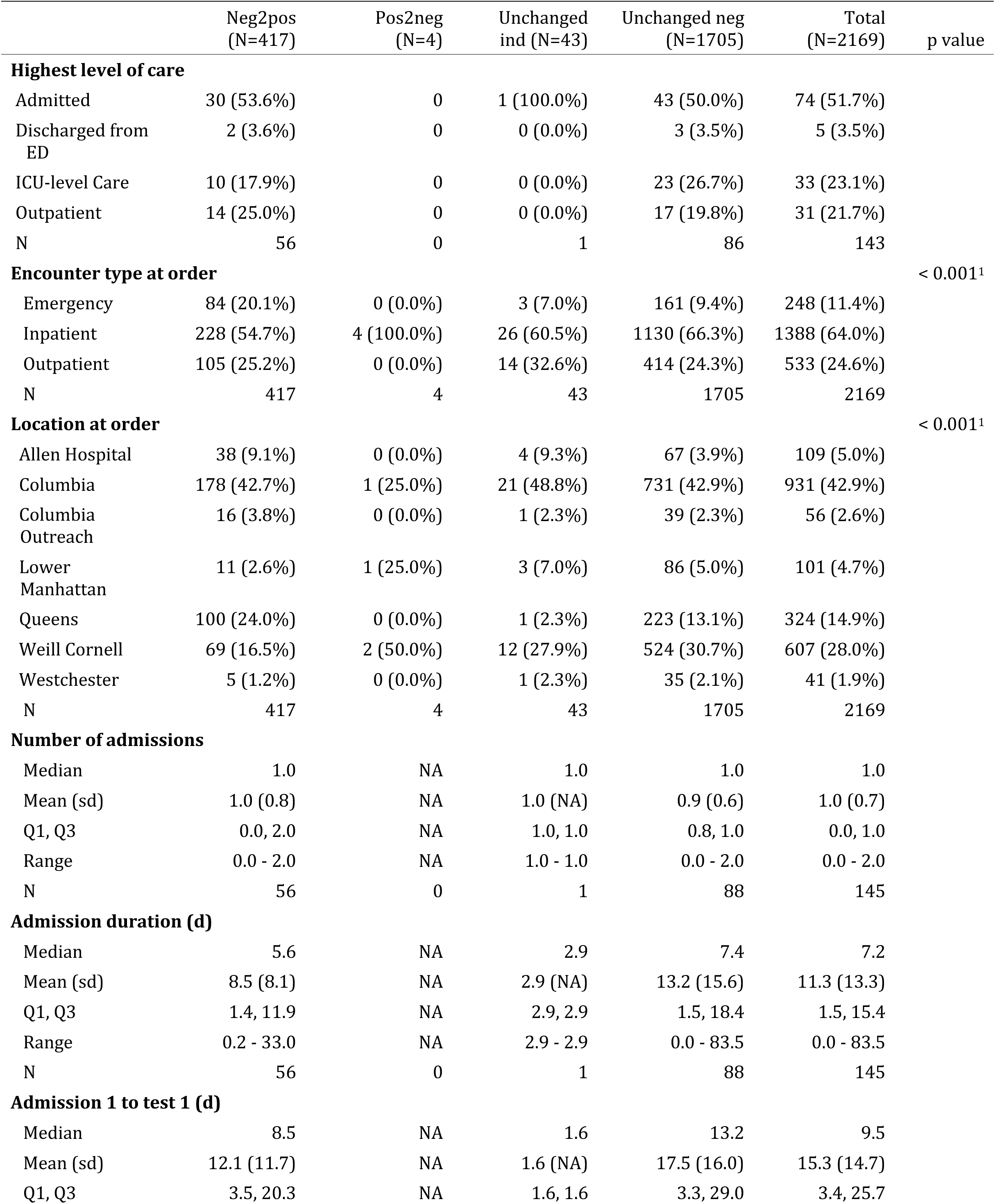
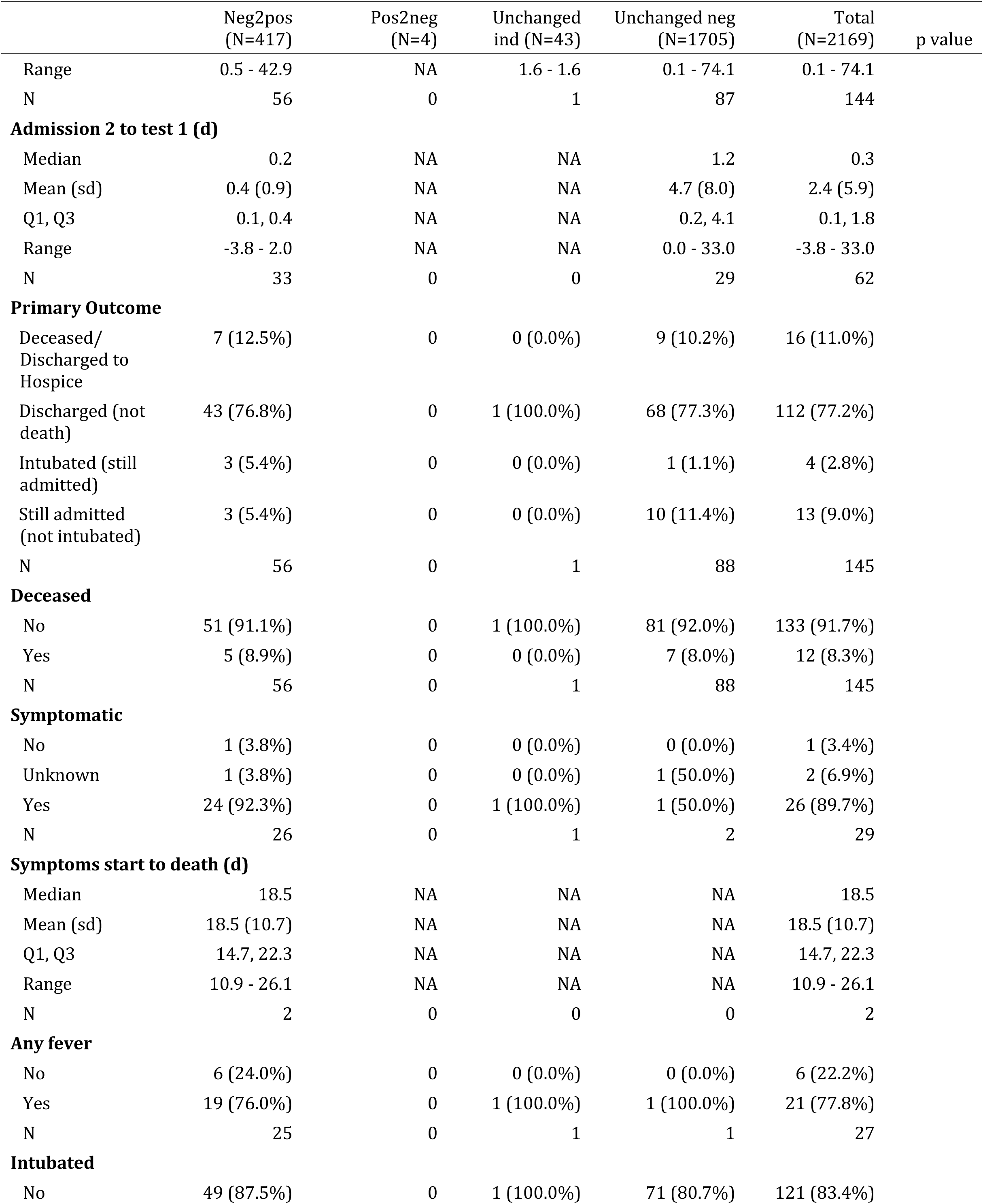
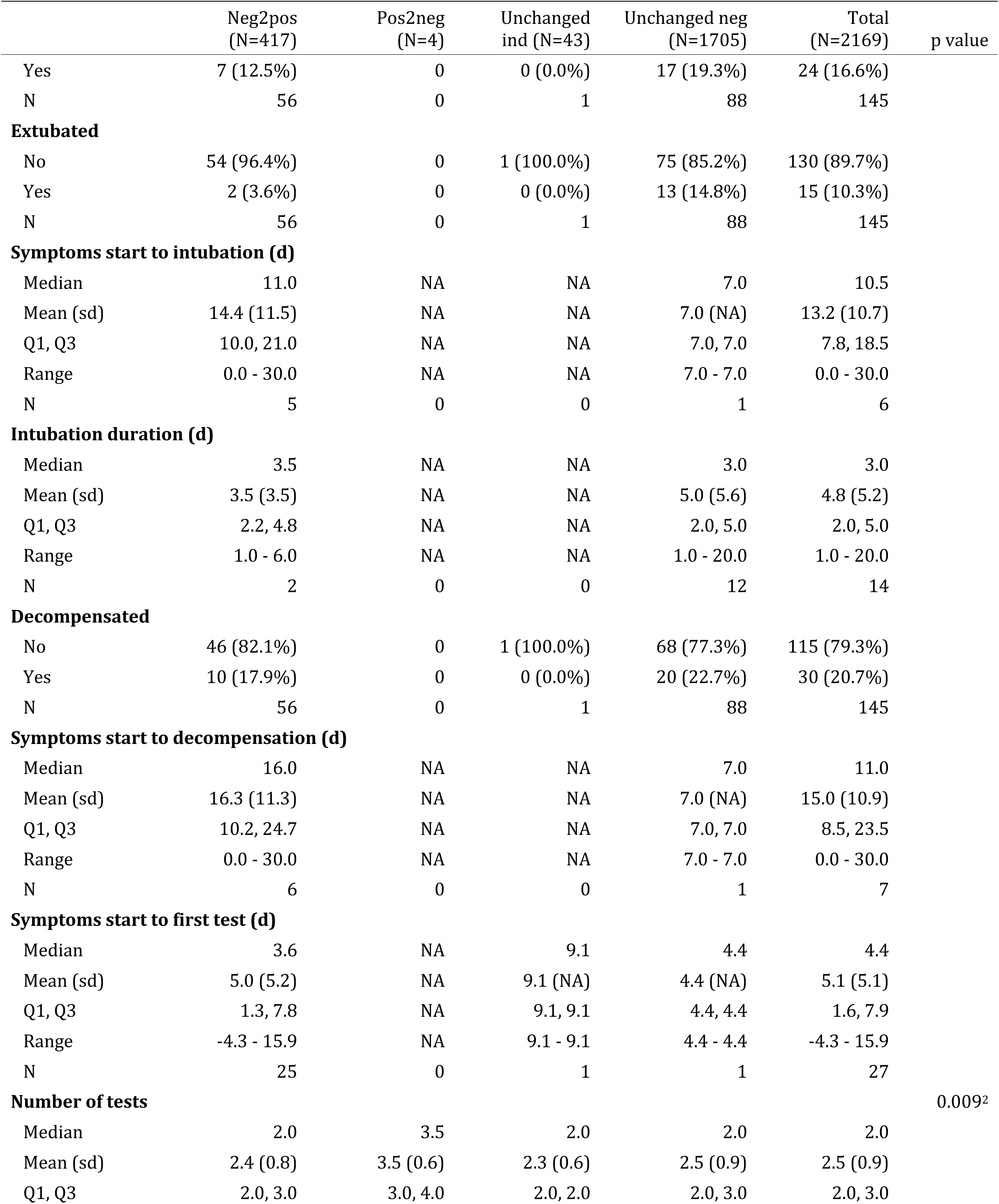
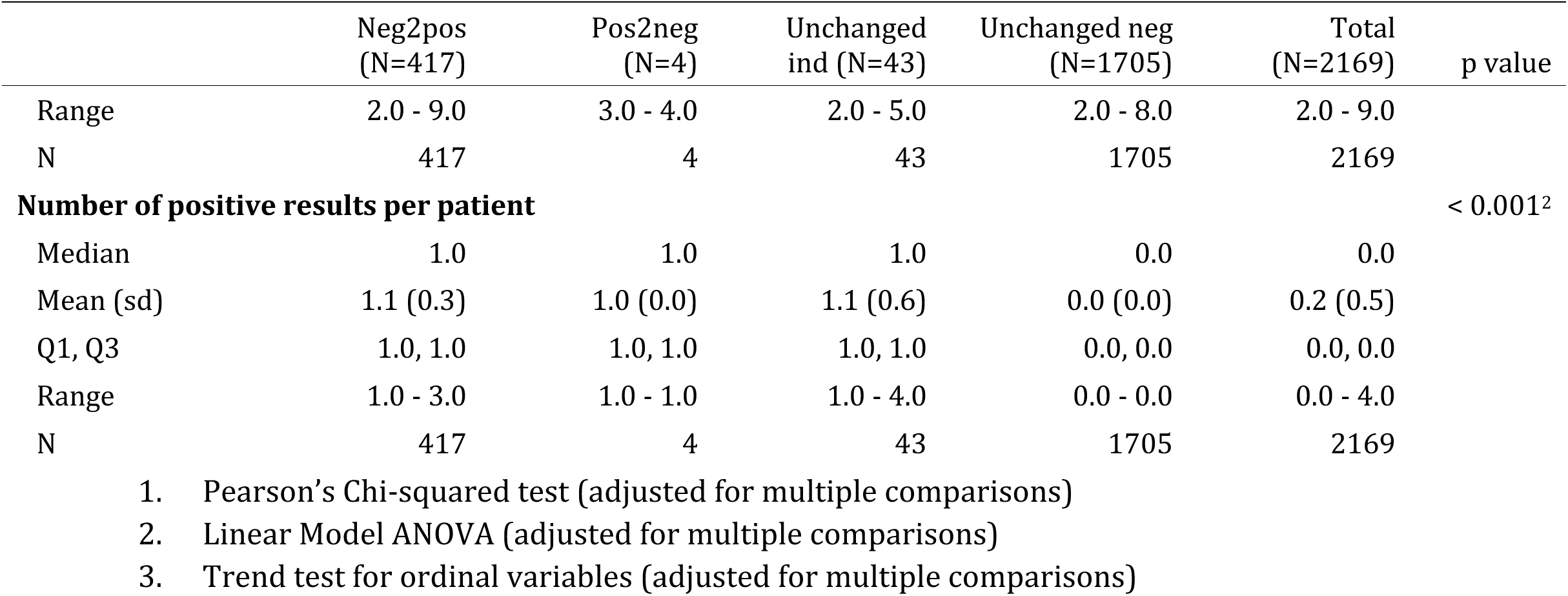
Characteristics of repeat-tested initially negative patients comparing those that converted from negative to positive (Neg2pos), with those that remained negative (Unchanged neg) or indeterminate (Unchanged ind) patients.

